# Using routinely-collected, linked data to measure and monitor health system performance in Australia: development of an indicator of continuity of primary health care

**DOI:** 10.1101/2024.12.10.24318818

**Authors:** Jennifer Welsh, Kay Soga, Rachel Freeman Robinson, Danielle Butler, Rachael Eddowes, Amelia Yazidjoglou, Angus Douglas, Kirsty Douglas, Nina Lazarevic, Hsei-Di Law, Grace Joshy, Sue Trevenar, Tsheten Tsheten, Adrian Webster, Michael Frost, Bernice Cropper, Sally Hall Dykgraaf, Christine Phillips, Emily Banks, Rosemary J Korda

**Author notes:** **Ethics approval** Ethics approval for this study was granted by the Australian National University Human Research Ethics Committee (HREC 2021/619). **Data availability** Data from the Person-Level Integrated data asset are available for approved projects to approved government and non-government users. Person Level Integrated Data Asset (PLIDA) | Australian Bureau of Statistics (abs.gov.au). **Declaration of funding** This project was funded by the Australian Institute of Health and Welfare and the Australian Government’s Medical Research Future Fund (grant number 2006309). Emily Banks receives support from the National Health and Medical Research Council (grant number 1136128). **Author contributions** JW, RK, EB and DB designed the study. JW, KS, RE, NL and HDL contributed to the analysis of the data. JW, AD, AY, RFR, DB and KD drafted the manuscript. All authors read and critically reviewed the manuscript.

## Abstract

**Objective:** To examine a potential new Australian health system indicator—continuity of care (CoC) in general practice—derived from whole-of-population linked administrative data.

**Methods:** Using data from Census 2016 and 2021 and the Medicare Benefits Schedule, available via the Person-Level Integrated Data Asset (PLIDA), we examined an indicator of CoC in general practice, the Usual Provider Index (UPI), over eight overlapping 2-year periods (2016-17 to 2022-23), and the 2-year period following the introduction of universal telehealth in April 2020). We measured population coverage of the UPI, sociodemographic- and health-related variation in high COC (UPI≥0.7), and changes in CoC over time.

**Results:** Population coverage of the UPI was high (>75%); higher for groups with greater healthcare needs, including older people and people with chronic conditions. Coverage decreased with increasing remoteness. In 2022-23, 36% of those with a UPI score had high CoC; proportions with high CoC were greater for older people, those born overseas, not proficient in English, and with health conditions, but lower for people living in more remote areas. Prior to 2020, 32-33% of the population had high CoC, increasing to 39% in the 2-year period directly following the introduction of universal telehealth.

**Conclusion:** When derived from whole-of population linked administrative data, the UPI is a suitable indicator of CoC for people with varying healthcare needs, and across different settings. Using new national data assets, the UPI could be included in routine health system reporting as one indicator of quality of care.

**Online Short Summary:** *What is known?:* Health system performance monitoring aims to facilitate health system planning and improvement. Whole-of-population linked administrative data may be used to extend current health system indicators in Australia.

*What this paper adds?:* We examined a potential new indicator—continuity of care within general practice—by assessing coverage and sensitivity to differences between groups and over time. Coverage was high, those with greater healthcare need had higher continuity, and continuity increased after the introduction of telehealth.

*What are the implications?:* New national data assets can be used to derive health system indicators, enhancing insight into Australian healthcare.

## Background

Measuring and monitoring health system performance aims to facilitate health system planning and improvement.^1,2^ Monitoring involves routine analysis of health system indicators, typically reflecting effectiveness, efficiency, quality and equity,^1,3^ to promote accountability, identify areas of strength or in need of reform, and evaluate the impact of policy change.^1,2,4^

In Australia, health system monitoring is governed at the national level by frameworks such as the Australian Health Performance Framework (hereafter: the Framework).^5^ The Framework includes indicators that are mostly outcome-based (e.g., incidence of ischaemic heart disease) rather than process-based (reflecting best practice care; e.g., proportion of women with an antenatal visit in the first trimester). While outcome-based indicators can identify problems, process-based indicators can facilitate insights into points of intervention.^5^

Advancements in the availability of routinely-collected, whole-of-population linked administrative data present opportunities to enhance indicators within Australia.^6^ One area with potential for improvement is primary care, which is fundamental to an effective and equitable health care system.^7^ Primary care is designed as a first point of access for patients, providing ongoing, coordinated care throughout the lifecourse,^8^ but only one of the core set of indicators within the Framework relates to primary care: bulk billing for non-referred general practitioner (GP) attendances.

One potential new indicator related to primary care is a measure of continuity of care (CoC), which refers to an ongoing patient-provider relationship.^9–11^ There are three types of CoC: informational (using information from past events to inform current care), management (consistent approach to managing conditions) and relational (an ongoing trusted relationship between patient and provider).^11^ Continuity of care is a core function of quality primary care,^7^ and has been shown to facilitate trust between patients and their GPs, improve coordination of services, and is associated with higher patient satisfaction,^9,12,13^ lower cost^12,14,15^ and better health outcomes.^16–19^ Thus, an indicator of continuity in primary care may provide insights into one aspect of quality of care that has been shown to support good patient outcomes.

Continuity of care can be measured using the Usual Provider Index (UPI). The UPI, a measure of relational CoC, calculates the proportion of visits to the most visited GP over a 2-year period. The UPI is potentially suitable for monitoring purposes as it can be readily calculated from routinely-collected health service data,^20–22^ including in Australia.^23^ Whether the UPI is a useful health system indicator is currently unclear and requires whole-of-population evidence on coverage and sensitivity to existing variation, including change from policy reform.

This paper aims to assess the utility of the UPI, derived from routinely-collected, whole-of-population linked administrative data, as an indicator of relational CoC in primary care, for health system monitoring in Australia. We do this by quantifying population coverage and variation in the UPI in relation to key characteristics and over time, including in relation to relevant policy change, for the whole of Australia. Based on available evidence, our *a priori* assumptions were that CoC will be higher among older patients,^24^ people of lower socioeconomic status,^25,26^ those with higher health care need,^25^ including those with chronic health conditions and in poorer health,^24^ and lower among people living in rural and remote areas.^26^ We expect to see little variation in continuity prior to 2020, but an increase in CoC from April 2020 when universal telehealth, which was conditional on an established patient-provider relationship, was introduced in Australia (supplementary material).

## Methods

We used data from the 2016 and 2021 Census of Population and Housing, Medicare Benefits Schedule (MBS), Death Registrations and migration data, available through the Person-Level Integrated Data Asset (PLIDA).^27^ We examined outcomes over eight 2-year overlapping study periods: 2016-17, 2017-18, 2018-19, 2019-20, 2020-21, 2021-22, 2022-23, and 1 April 2020-31 March 2022 (the period following the introduction of universal telehealth).

For each study period, we constructed a full study population and a (subset) CoC study population. Both study populations included Australian residents with a Census record that linked to the PLIDA Spine who were alive at the start of the study period, not overseas for the study period and had at least one out-of-hospital MBS claim for any service. The CoC study population additionally included those who had at least four GP visits over two years (supplementary material). Census 2016 was used to create study populations for each of the eight study periods; Census 2021 was used to create a study population for 2022-23.

Our outcome was CoC, measured with the UPI, derived from MBS claims data. For individuals with at least four GP visits in a two-year period, UPI was calculated,^29^ and we defined high CoC as a UPI score of ≥0.7 (i.e. minimum 70% of visits with one provider), consistent with previous research.^30^ We measured sociodemographic and health related characteristics (supplementary material), grouped into categories as shown in Table 1. Missing data were included as a separate category.

**Table 1.** Study population numbers Proportion of the Census 2021 study population with at least 4 GP visits in 2022-23, by sociodemographic- and health-related characteristics.

|  | ≥ 4 GP Visits (CoC study population) | Census 2021 full study population | Proportion ≥ 4 GP visits (UPI coverage) |
| --- | --- | --- | --- |
| Total | 17,322,272 | 22,540,630 | 76.8 |
| Sex |  |  |  |
| Male | 7,775,807 | 11,009,562 | 70.6 |
| Female | 9,546,465 | 11,531,068 | 82.8 |
| Age Group (years) |  |  |  |
| 0-14 | 2,525,838 | 4,125,370 | 61.2 |
| 15-24 | 1,756,017 | 2,609,887 | 67.3 |
| 25-44 | 4,493,042 | 6,038,455 | 74.4 |
| 45-69 | 5,856,056 | 6,921,805 | 84.6 |
| 70+ | 2,691,319 | 2,845,113 | 94.6 |
| Relationship status |  |  |  |
| Married/de facto | 8,553,219 | 10,330,681 | 82.8 |
| Single | 4,955,563 | 6,299,860 | 78.7 |
| Missing | 692,000 | 860,794 | 80.4 |
| Employment status |  |  |  |
| Employed | 8,119,560 | 10,651,014 | 76.2 |
| Unemployed | 409,239 | 562,017 | 72.8 |
| Not in the labour force | 2,406,582 | 3,035,692 | 79.3 |
| Missing | 79,805 | 106,506 | 74.9 |
| Education Level |  |  |  |
| University | 3,834,398 | 4,740,615 | 80.9 |
| High school and/or other qualification | 5,826,486 | 7,173,754 | 81.2 |
| No qualification | 3,084,122 | 3,533,040 | 87.3 |
| Missing | 221,419 | 253,704 | 87.3 |
| Remoteness area |  |  |  |
| Major cities | 12,447,415 | 15,920,363 | 78.2 |
| Inner regional | 2,891,212 | 3,841,441 | 75.3 |
| Outer regional | 1,158,130 | 1,620,995 | 71.4 |
| Remote | 125,559 | 196,521 | 63.9 |
| Very remote | 48,091 | 76,318 | 63.0 |
| Other | 651,865 | 884,992 | 73.7 |
| Country of birth |  |  |  |
| Aus/NZ | 12,651,344 | 16,819,619 | 75.2 |
| Other Oceania | 110,844 | 138,045 | 80.3 |
| North-West Europe | 1,135,751 | 1,368,689 | 83.0 |
| Southern and Eastern Europe | 538,092 | 607,171 | 88.6 |
| North Africa and Middle East | 357,446 | 413,965 | 86.3 |
| South-East Asia | 669,594 | 872,364 | 76.8 |
| North East Asia | 494,084 | 655,055 | 75.4 |
| Southern and Central Asia | 704,200 | 842,763 | 83.6 |
| Americas | 210,281 | 262,680 | 80.1 |
| Sub-Saharan Africa | 264,897 | 336,651 | 78.7 |
| Missing | 185,739 | 223,628 | 83.1 |
| English language proficiency |  |  |  |
| Proficient | 13,613,435 | 16,818,577 | 80.9 |
| Speaks English not well or not at all | 492,480 | 560,432 | 87.9 |
| Missing | 94,867 | 112,326 | 84.5 |
| SEIFA IRSD |  |  |  |
| Most disadvantaged | 3,128,218 | 4,044,670 | 77.3 |
| Quintile 2 | 3,271,149 | 4,242,191 | 77.1 |
| Quintile 3 | 3,377,039 | 4,388,957 | 76.9 |
| Quintile 4 | 3,435,712 | 4,468,599 | 76.9 |
| Least disadvantaged | 3,435,294 | 4,481,311 | 76.7 |
| Missing | 674,860 | 914,902 | 73.8 |
| Household income |  |  |  |
| \$104K+ | 2,708,714 | 3,589,174 | 75.5 |
| \$65K-<\$104K | 4,091,000 | 5,416,491 | 75.5 |
| \$26K-\$65K | 6,500,051 | 8,463,220 | 76.8 |
| \$1-<\$26K | 2,292,620 | 2,829,458 | 81.0 |
| Other (including missing) | 1,729,887 | 2,242,287 | 77.1 |
| Health conditions |  |  |  |
| Arthritis | 1,932,148 | 2,056,873 | 93.9 |
| Asthma | 1,697,665 | 1,989,278 | 85.3 |
| Cancer | 639,861 | 689,370 | 92.8 |
| Dementia | 148,384 | 161,282 | 92.0 |
| Diabetes | 1,081,524 | 1,130,911 | 95.6 |
| Heart disease | 889,897 | 940,085 | 94.7 |
| Kidney disease | 198,939 | 213,912 | 93.0 |
| Lung disease | 385,475 | 412,717 | 93.4 |
| Mental health condition | 1,923,348 | 2,140,207 | 89.9 |
| Stroke | 204,348 | 217,150 | 94.1 |
| Number of named conditions |  |  |  |
| None of the conditions | 11,260,992 | 15,810,292 | 71.2 |
| 1 condition | 4,036,828 | 4,584,057 | 88.1 |
| 2 conditions | 1,339,709 | 1,424,248 | 94.1 |
| 3 or more conditions | 684,743 | 722,033 | 94.8 |

Notes: English language proficiency and marital status are measured for people aged 18 years and over at the time of Census 2021. Results for employment status are reported for people aged 15-64 years at the time of Census 2021. Results for highest level of education are reported for people aged 25 years and older.

To quantify population coverage of the UPI, we reported proportions of the full study population with at least four GP visits in each 2-year period. We focused our analysis on the most recent study period (2022-23) using the full Census 2021 study population as the denominator. We also examined the extent to which UPI coverage changed over time by describing proportions of the full Census 2016 study population with at least 4 GP visits in each study period.

To examine socio-demographic and health-related variation in high CoC, we estimated proportions of the Census 2021 CoC study population with high CoC in 2022-23. To examine whether variation in high CoC could be accounted for by other characteristics, we estimated prevalence ratios (PRs) and 95% confidence intervals (CIs) using Poisson regression with robust standard errors.^33^ For each set of exposure-outcome associations, we ran three models: i) unadjusted, ii) adjusted for sex and 5-year age group (0-4 to 80+), and iii) additionally adjusted for remoteness. Associations between sociodemographic characteristics and high CoC were further adjusted for presence of the specified health conditions (none/≥one; model 4).

We examined temporal trends in proportion of Census 2016 CoC study population with high CoC. To examine whether the UPI was sensitive to policy-related change, we compared trends in the proportions with high CoC during the pandemic period (Apr-2020–Mar2022, and 2021-22, 2022-23) to pre-pandemic periods.

We assessed potential for selection bias introduced by restricting our study populations to those with a linked Census record, by comparing population coverage and proportions with high CoC between our Census 2016 study populations and all records in MBS data. For these analyses, the full MBS study population was people with at least 1 MBS claim in the study period, and the MBS CoC study population was those in the MBS study population with at least 4 GP visits. We assessed the sensitivity of our findings to our definition of high CoC (UPI≥0.7) by describing proportions of the population with high CoC using alternative cut points (0.75, 0.80). To investigate whether presence of a health condition accounted for sociodemographic variation in high CoC, we estimated PRs among people with at least one of the measured health conditions. We also estimated descriptive statistics for continuous UPI scores in each study period.

Ethics approval for this study was granted by the Australian National University Human Research Ethics Committee (HREC 2021/619).

## Results

After exclusions (n=2.9M, 11.5%), there were 22,540,630 people with a Census 2021 record and at least one MBS claim in the 2022-23 study period (Supplementary Figure 1). Study population numbers were similar in other study periods (Supplementary Figure 2).

Of the Census 2021 study population, 77% (n=17,322,272) had ≥4 GP visits in 2022-23 and were eligible for a UPI score (Table 1). Coverage of the UPI increased with age and was higher for women compared to men, people not in the labour force compared to employed people, people with lower levels of education compared to high education, those not proficient in English compared to proficient English speakers, and those born in Southern and Eastern Europe or North Africa and the Middle East compared to Australia or New Zealand (Table 1). Coverage was lower with increasing remoteness and while there were little differences by SEIFA IRSD, coverage was higher for those with lower-compared to higher-household income. Overall, UPI coverage was high for people with chronic health conditions and increased with more conditions.

Coverage and sociodemographic variation in coverage did not substantially change over study periods between 2016 and 2022 (Supplementary Table 2, Supplementary Figure 3).

In 2022-23, 36.0% of the Census 2021 CoC study population had high CoC (Supplementary Table 3, Supplementary Figure 4). Proportions with high CoC increased with age and were higher for men compared to women, those not in the labour force or unemployed compared to those employed, those with lower levels of education, people born overseas and those not proficient in English compared to those who were (Figure 1). Proportions with high CoC were also higher among those living in more disadvantaged areas and in households with lower incomes. Proportions with high CoC also increased with increasing number of GP visits, and among those with chronic health conditions, except for asthma, and decreased with increasing remoteness.

**Figure 1.**
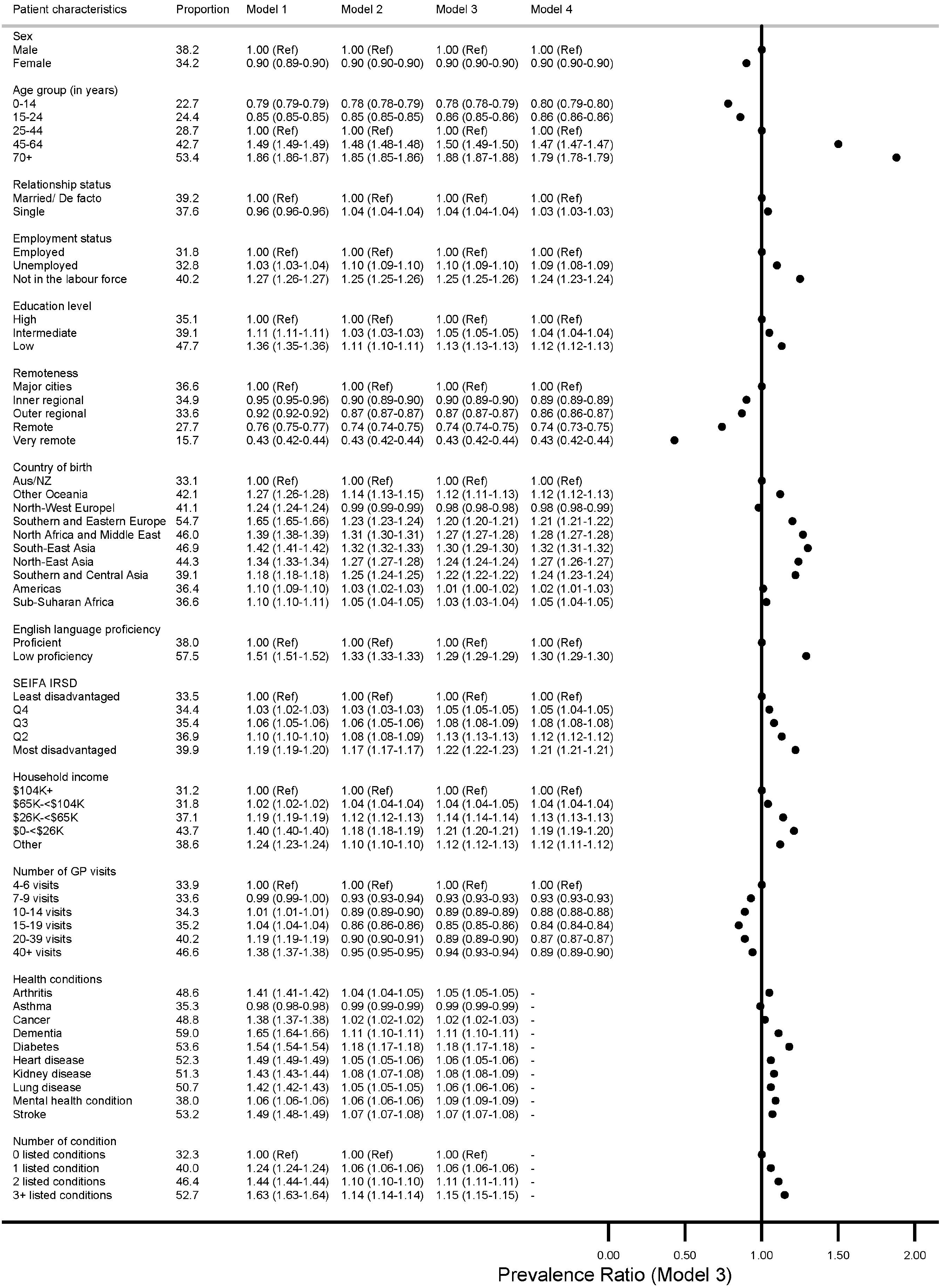
Proportion of the Census 2021 continuity of care study population with high continuity of care and prevalence ratios (and 95% confidence intervals) describing the association between high continuity and socio-demographic and health characteristics, 2022-23.

Adjustment for age group, sex and remoteness attenuated the associations between high CoC and sociodemographic- and health-related factors (Figure 1, models 2 and 3) but did not fully account for the associations. Further adjustment for health condition did not materially change the relationships (Figure 1, model 4). While the absolute proportions of people with high CoC increased when restricted to people with at least one health condition, sociodemographic variation was similar to the full study population (Supplementary Figure 5).

Prior to 2020, 32-33% of the Census 2016 CoC study population had high CoC (Figure 2). This increased to 39.1% in the 2 years following the introduction of telehealth and then reduced slightly to 37.2% in 2022-23. An increase was observed across all sociodemographic groups but was less pronounced in groups with a higher proportion of high CoC prior to the pandemic (Supplementary Table 3, Supplementary Figure 4).

**Figure 2.**
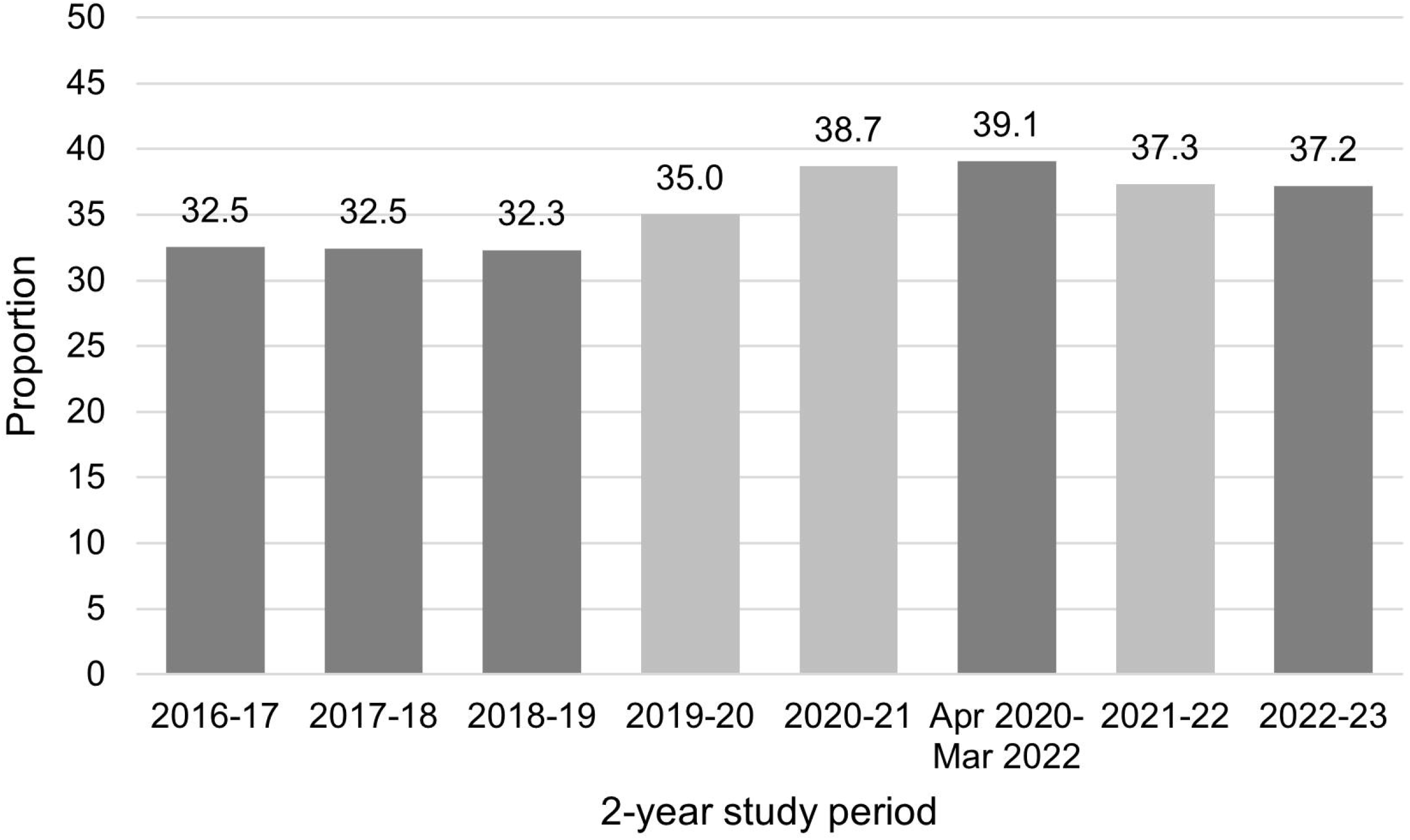
Proportion of the Census 2016 continuity of care study population with high continuity of care over the study period. Figure 2 note: High continuity of care is defined as a usual provider index score of ≥0.7. Grey bars contain data that is included in the period 1 April 2020-31 March 2022. Proportions with high continuity by sociodemographic characteristics are provided in Supplementary Table 3.

Proportions of people with high CoC were generally 1-2 percentage points higher in the Census 2016 study populations relative to the MBS study population (Supplementary Tables 4 and 5). Differences were larger for younger people (up to three percentage points, likely reflecting cohort effects within the Census study population) and those with missing location data (up to 11 percentage points) but sociodemographic variation was essentially unchanged. Variation in high CoC did not differ materially when using different cut points of high CoC (Supplementary Table 6) and variation in continuous UPI scores mirrored patterns observed using the dichotomous (high vs other) variable (Supplementary Table 7).

## Discussion

This study has shown that the UPI could be derived for >75% of the Australian population and coverage was generally higher among populations with greater healthcare need. Consistent with our *a priori* expectations, proportions with high CoC increased with age, and was higher for people with chronic health conditions, those living in lower income households, people of lower socioeconomic position, and people living in more urban areas. Around one third of the CoC population had high CoC prior to the pandemic, which increased by around five percentage points after the introduction of telehealth. While it is unclear whether this increase was a direct result of the policy change (which required an established patient-provider relationship), particularly given that there were several changes to the health care system during the pandemic, this finding is also consistent with our *a priori* assumptions and serves as preliminary evidence that the UPI is sensitive to changes over time.

Our findings suggest that the UPI is a useful indicator of health system performance and could be included in routine health reporting in Australia. This would add a process-based primary care indicator to existing indicators, providing nuanced insights in performance while highlighting opportunities for reform. Our finding that high CoC was higher for several priority populations highlights one area in which the health system is working to improve equity but that most Australians did not have high CoC also indicates opportunities for improvement. Ongoing monitoring of CoC, including equity in CoC, can support preservation of high CoC where it exists and increase CoC where required. Additional analyses examining variation within and across Primary Healthcare Networks (PHNs) —where actionable interventions could occur—may facilitate interventions, and are also possible with these linked data.

Although our findings suggest that the UPI is broadly suitable for routine monitoring, it does have limitations. First, the UPI can only be derived for people with a minimum number of visits over a specified period.^21^ Interpretation of findings should be done in reference to the proportion with a UPI score. Second, given the measure calculates the proportion of visits with the usual provider, it may be sensitive to the underlying number of GP visits. While we saw no evidence that using different cut points to define high CoC altered the pattern of results, differences between populations in use of GP services should be considered. Third, while high continuity is associated with quality care and good patient outcomes, it is only an indicator and not a guarantee of quality care. Fourth, the UPI is a measure of relational CoC and does not measure other aspects of CoC when care is delivered across multiple practitioners within a single practice.^21^ Practice identifiers are not currently available within MBS data, although there have been efforts to create proxy identifiers.^34^ Including these data within existing national data assets would enable indicators of CoC at the practice level, and could be used to understand the types of practices that do and do not support CoC, as well as improve understanding of the interplay between relational (same provider) and informational and management (same practice) CoC. Lastly, the UPI provides a proxy measure for relational CoC that can be measured with existing administrative data for the whole population. It is unable to capture other critical aspects of this construct, such as sense of trust and depth of knowledge of the patient’s history and context, which is integral to how CoC contributes to improved outcomes.

This study provides insights into the benefits and methodological challenges associated with using national data assets for monitoring purposes. While CoC can be derived using unlinked MBS data, linking MBS to Census, which contains individual- and household-level information not available in MBS, could substantially improve reporting of equity in health system indicators. However, linked data can introduce bias given incomplete census response^35^ and linkage error, both of which are likely to be differential for priority groups compared to the general population. Furthermore, given that Census data are collected every five years, using these data to report on indicators in intercensal years excludes new arrivals (babies, new migrants) and may include out-of-date information on time-varying characteristics (e.g. employment status). We observed small differences in estimates when using the MBS-compared to the Census population, and between the two Census study populations for overlapping periods. While differences were small and conclusions substantially unchanged, these issues should be considered, particularly when examining long-term trends.

Routinely-collected linked administrative data can be used to derive new indicators of health system performance, including measures that capture process-based indicators such as CoC, one aspect of quality of care. These data provide opportunities to enhance health system performance to support an effective, equitable and sustainable health system.

## Data Availability

Data from the Person-Level Integrated data asset are available for approved projects to approved government and non-government users.

https://www.abs.gov.au/about/data-services/data-integration/integrated-data/person-level-integrated-data-asset-plida

## Acknowledgements

We acknowledge the contributions of the Study Steering Committee members and members of the consumer engagement team.

