## Supplementary Material for "Using routinely-collected, linked data to measure and monitor health system performance in Australia: development of an indicator of continuity of primary health care"

### Additional information on Methods

#### Linkage of data sources with the Person Level Integrated Data Asset (PLIDA)

Census, Death Registrations and migration records were linked via the PLIDA Spine^1^ using deterministic linkage methods, based on name, date of birth, address and sex. A direct link exists between the Spine and MBS data.

#### Variables used in analyses

We measured the following variables: sex, age group (based on age at 1 January in each study period), English language proficiency (for ≥18years), marital status (≥18years), employment status (15-64years), highest level of education (≥25years), equivalised household income, remoteness, country of birth,^2^ Socio-Economic Indexes for Areas Index of Relative Socio-economic Disadvantage (SEIFA IRSD)^3^ in population-based quintiles, and frequency of GP visits in the two-year study period. For analyses using Census 2021, we also measured self-reported number of health conditions which was also used to derive number of specified conditions (0, 1, 2, ≥3). The heal conditions included: arthritis, asthma, cancer (including remission), dementia (including Alzheimer's disease), diabetes (excluding gestational diabetes), heart disease (including heart attack or angina), kidney disease, lung disease (including COPD or emphysema), mental health condition (including depression or anxiety) and stroke. Information on health conditions were not collected on Census 2016. These were ascertained from the Census, except for sex and age (ascertained from Medicare Consumer Directory), remoteness and SEIFA IRSD (derived from address recorded within the Medicare Consumer Directory on the 31 January of each study period), and frequency of GP visits (derived from MBS claims data).

#### MBS-funded reimbursements for these GP services

From 13 March 2020, GPs were able to claim reimbursement for services provided under Medicare for telehealth appointments (prior to this, telehealth had been largely restricted to people living in remote areas). This policy was in response to the COVID-19 pandemic and was designed to reduce risk of community transmission (including to health care providers) while maintaining health care. These MBS items were dependent on the GP having an established clinical relationship with the patient, which was defined as:

1. The provider having seen the patient face to face in the last 12 months or
2. The provider was providing services through a practice that the patient had attended face to face in previous 12 months.

More information can be found here: [MBS Online - COVID-19 Temporary MBS Telehealth Services](https://www.mbsonline.gov.au/internet/mbsonline/publishing.nsf/Content/Factsheet-TempBB)

#### Definition of GP visits

For all analyses, we excluded in-hospital MBS service claims and invalid MBS claims. The definition of ‘GP visit’ is a unique combination of synthetic ID, date of service, and service provider ID in the linked dataset. Under this definition, a patient who saw two different GPs on the same day had two ‘GP visits.’

### Supplementary tables

#### Supplementary Table 1: MBS codes used for GP consultations^1^

| **In-person** | **Telephone (audio-only)** | **Video** |
| --- | --- | --- |
| **All item numbers from following MBS groups and subgroups^2^ :** | | |
| A1, A2, A6, A7.2, A7.4, A7.5, A7.6, A7.7, A7.8, A7.9, A7.10, A7.11, A11, A14, A15, A17, A18, A19, A20, A22, A23, A27, A35, A36.1, A36.4, A45.1 | A40.2, A40.10, A40.12, A40.14, A40.16, A40.20, A40.22, A40.26, A40.28, A40.30, A40.40, A45.3 | A7.12, A30, A40.1, A40.3, A40.11, A40.13, A40.15, A40.19, A40.21, A40.25, A40.27, A40.39, A45.2 |
| **Additional specific item numbers^3^** | | |
| 91283, 91285, 91286, 91287, 91721, 91723, 91725, 91727 |  | 91371, 91372, 91729, 91731 |
| 93287, 93288, 93300, 93303, 93291, 93292, 93306, 93309, 93400, 93401, 93402, 93403, 93421, 93431, 93432, 93433, 93434, 93451, 93469, 93470, 93475, 93479, 93715 | 92746, 92747, 93302, 93305, 93308, 93311, 93408, 93409, 93410, 93411, 93423, 93439, 93440, 93441, 93442, 93453, 93716, 93717 | 93301, 93304, 93307, 93310, 93404, 93405, 93406, 93407, 93422, 93435, 93436, 93437, 93438, 93452 |

1. All items are those claimed by general practitioners or other medical practitioners (not including specialist or consultant physician) providing primary care services

2. MBS group A44 was excluded because these pertained solely to delivery of COVID-19 vaccines and do not reflect usual general practice MBS services.

3. Additional specific items were used where the MBS group or subgroup were not homogenous in modality.

#### Supplementary Table 2. Study population numbers and proportion of the Census 2016 study population with at least 4 GP visits in each of the 2-year study periods, by sociodemographic characteristics

|  | 2016-17 | | | 2017-18 | | | 2018-19 | | | 2019-20 | | | 2020-21 | | | 2021-22 | | | 2022-23 | | |
| --- | --- | --- | --- | --- | --- | --- | --- | --- | --- | --- | --- | --- | --- | --- | --- | --- | --- | --- | --- | --- | --- |
|  | ≥4 GP visits (‘000) | Study pop (‘000) | % | ≥4 GP visits (‘000) | Study pop (‘000) | % | ≥4 GP visits (‘000) | Study pop (‘000) | % | ≥4 GP visits (‘000) | Study pop (‘000) | % | ≥4 GP visits (‘000) | Study pop (‘000) | % | ≥4 GP visits (‘000) | Study pop (‘000) | % | ≥4 GP visits (‘000) | Study pop (‘000) | % |
| Total | 15,943 | 19,708 | 81 | 15,907 | 19,664 | 81 | 15,906 | 19,537 | 81 | 15,744 | 19,376 | 81 | 15,364 | 19,364 | 79 | 15,233 | 19,271 | 79 | 15,069 | 18,934 | 80 |
| Sex |  |  |  |  |  |  |  |  |  |  |  |  |  |  |  |  |  |  |  |  |  |
| Male | 7,198 | 9,499 | 76 | 7,181 | 9,477 | 76 | 7,188 | 9,414 | 76 | 7,110 | 9,334 | 76 | 6,907 | 9,372 | 74 | 6,827 | 9,328 | 73 | 6,718 | 9,111 | 74 |
| Female | 8,746 | 10,208 | 86 | 8,727 | 10,188 | 86 | 8,718 | 10,123 | 86 | 8,634 | 10,042 | 86 | 8,457 | 9,993 | 85 | 8,407 | 9,943 | 85 | 8,351 | 9,824 | 85 |
| Age Group (years) | |  |  |  |  |  |  |  |  |  |  |  |  |  |  |  |  |  |  |  |  |
| 0-14 | 2,784 | 3,912 | 71 | 2,533 | 3,662 | 69 | 2,344 | 3,416 | 69 | 2,090 | 3,157 | 66 | 1,697 | 2,911 | 58 | 1,502 | 2,671 | 56 | 1,379 | 2,388 | 58 |
| 15-24 | 1,758 | 2,346 | 75 | 1,758 | 2,346 | 75 | 1,768 | 2,344 | 75 | 1,751 | 2,341 | 75 | 1,711 | 2,373 | 72 | 1,701 | 2,389 | 71 | 1,667 | 2,360 | 71 |
| 25-44 | 4,203 | 5,334 | 79 | 4,196 | 5,300 | 79 | 4,203 | 5,263 | 80 | 4,170 | 5,224 | 80 | 4,094 | 5,246 | 78 | 4,043 | 5,235 | 77 | 3,956 | 5,134 | 77 |
| 45-69 | 5,234 | 6,064 | 86 | 5,320 | 6,151 | 86 | 5,388 | 6,205 | 87 | 5,432 | 6,244 | 87 | 5,466 | 6,322 | 86 | 5,487 | 6,359 | 86 | 5,482 | 6,348 | 86 |
| 70+ | 1,964 | 2,051 | 96 | 2,101 | 2,205 | 95 | 2,203 | 2,309 | 95 | 2,301 | 2,410 | 96 | 2,397 | 2,511 | 95 | 2,501 | 2,616 | 96 | 2,585 | 2,705 | 96 |
| Relationship status |  |  |  |  |  |  |  |  |  |  |  |  |  |  |  |  |  |  |  |  |  |
| Married/de facto | 7,551 | 8,970 | 84 | 7,604 | 8,973 | 85 | 7,639 | 8,933 | 86 | 7,625 | 8,877 | 86 | 7,570 | 8,873 | 85 | 7,532 | 8,827 | 85 | 7,458 | 8,706 | 86 |
| Single | 4,398 | 5,299 | 83 | 4,408 | 5,293 | 83 | 4,410 | 5,258 | 84 | 4,372 | 5,209 | 84 | 4,312 | 5,211 | 83 | 4,262 | 5,172 | 82 | 4,169 | 5,052 | 83 |
| Missing | 806 | 953 | 85 | 785 | 928 | 85 | 754 | 888 | 85 | 730 | 858 | 85 | 707 | 841 | 84 | 687 | 822 | 84 | 666 | 796 | 84 |
| Employment status |  |  |  |  |  |  |  |  |  |  |  |  |  |  |  |  |  |  |  |  |  |
| Employed | 7,256 | 9,169 | 79 | 7,349 | 9,190 | 80 | 7,444 | 9,193 | 81 | 7,474 | 9,180 | 81 | 7,449 | 9,258 | 80 | 7,445 | 9,265 | 80 | 7,395 | 9,155 | 81 |
| Unemployed | 513 | 648 | 79 | 520 | 650 | 80 | 527 | 650 | 81 | 528 | 648 | 81 | 525 | 654 | 80 | 522 | 654 | 80 | 513 | 643 | 80 |
| Not in the labour force | 2,369 | 2,825 | 84 | 2,375 | 2,821 | 84 | 2,388 | 2,811 | 85 | 2,378 | 2,794 | 85 | 2,359 | 2,803 | 84 | 2,351 | 2,796 | 84 | 2,319 | 2,754 | 84 |
| Missing | 69 | 84 | 82 | 69 | 84 | 83 | 69 | 83 | 83 | 69 | 82 | 83 | 68 | 82 | 82 | 67 | 82 | 82 | 65 | 80 | 82 |
| Education Level |  |  |  |  |  |  |  |  |  |  |  |  |  |  |  |  |  |  |  |  |  |
| University | 2,808 | 3,484 | 81 | 2,840 | 3,497 | 81 | 2,873 | 3,493 | 82 | 2,877 | 3,480 | 83 | 2,865 | 3,489 | 82 | 2,879 | 3,488 | 83 | 2,878 | 3,454 | 83 |
| High school and/or other qualification | 5,225 | 6,236 | 84 | 5,251 | 6,228 | 84 | 5,268 | 6,196 | 85 | 5,254 | 6,154 | 85 | 5,213 | 6,158 | 85 | 5,170 | 6,124 | 84 | 5,100 | 6,024 | 85 |
| No qualification | 3,178 | 3,544 | 90 | 3,163 | 3,521 | 90 | 3,121 | 3,459 | 90 | 3,073 | 3,396 | 90 | 3,018 | 3,352 | 90 | 2,958 | 3,290 | 90 | 2,880 | 3,202 | 90 |
| Missing | 303 | 331 | 92 | 291 | 319 | 91 | 273 | 298 | 91 | 257 | 281 | 91 | 243 | 267 | 91 | 230 | 254 | 91 | 217 | 239 | 91 |
| Remoteness area |  |  |  |  |  |  |  |  |  |  |  |  |  |  |  |  |  |  |  |  |  |
| Major cities | 11,534 | 14,089 | 82 | 11,510 | 14,065 | 82 | 11,524 | 13,982 | 82 | 11,410 | 13,876 | 82 | 11,112 | 13,869 | 80 | 11,035 | 13,815 | 80 | 10,926 | 13,588 | 80 |
| Inner regional | 2,877 | 3,622 | 79 | 2,871 | 3,610 | 80 | 2,862 | 3,581 | 80 | 2,830 | 3,545 | 80 | 2,778 | 3,544 | 78 | 2,749 | 3,519 | 78 | 2,715 | 3,448 | 79 |
| Outer regional | 1,275 | 1,635 | 78 | 1,268 | 1,630 | 78 | 1,262 | 1,617 | 78 | 1,248 | 1,600 | 78 | 1,224 | 1,598 | 77 | 1,204 | 1,586 | 76 | 1,185 | 1,553 | 76 |
| Remote | 156 | 214 | 73 | 156 | 213 | 73 | 155 | 212 | 73 | 154 | 210 | 73 | 151 | 210 | 72 | 148 | 208 | 71 | 146 | 205 | 71 |
| Very remote | 80 | 121 | 66 | 81 | 120 | 68 | 81 | 119 | 68 | 80 | 118 | 68 | 79 | 118 | 67 | 77 | 117 | 66 | 77 | 115 | 67 |
| Other | 22 | 27 | 80 | 22 | 27 | 81 | 22 | 26 | 82 | 21 | 26 | 82 | 21 | 26 | 81 | 21 | 26 | 80 | 21 | 26 | 80 |
| Country of birth |  |  |  |  |  |  |  |  |  |  |  |  |  |  |  |  |  |  |  |  |  |
| Aus/NZ | 11,757 | 14,735 | 80 | 11,702 | 14,679 | 80 | 11,688 | 14,579 | 80 | 11,556 | 14,463 | 80 | 11,232 | 14,467 | 78 | 11,119 | 14,402 | 77 | 11,020 | 14,152 | 78 |
| Other Oceania | 102 | 121 | 85 | 102 | 120 | 85 | 102 | 119 | 85 | 102 | 119 | 86 | 100 | 119 | 84 | 99 | 118 | 84 | 98 | 116 | 84 |
| North-West Europe | 1,091 | 1,298 | 84 | 1,090 | 1,289 | 85 | 1,083 | 1,267 | 85 | 1,071 | 1,245 | 86 | 1,055 | 1,230 | 86 | 1,039 | 1,212 | 86 | 1,017 | 1,184 | 86 |
| Southern and Eastern Europe | 531 | 586 | 91 | 528 | 582 | 91 | 519 | 570 | 91 | 508 | 557 | 91 | 497 | 547 | 91 | 487 | 535 | 91 | 471 | 518 | 91 |
| North Africa and Middle East | 293 | 328 | 89 | 292 | 328 | 89 | 291 | 326 | 89 | 289 | 324 | 89 | 286 | 323 | 89 | 284 | 321 | 89 | 280 | 316 | 89 |
| South-East Asia | 555 | 688 | 81 | 559 | 693 | 81 | 568 | 695 | 82 | 567 | 695 | 82 | 562 | 701 | 80 | 566 | 704 | 80 | 563 | 695 | 81 |
| Northeast Asia | 392 | 506 | 77 | 403 | 517 | 78 | 415 | 524 | 79 | 412 | 524 | 79 | 406 | 527 | 77 | 415 | 530 | 78 | 412 | 525 | 79 |
| Southern and Central Asia | 521 | 607 | 86 | 532 | 619 | 86 | 543 | 628 | 87 | 547 | 631 | 87 | 545 | 637 | 86 | 551 | 642 | 86 | 547 | 636 | 86 |
| Americas | 167 | 206 | 81 | 170 | 208 | 82 | 172 | 208 | 83 | 173 | 208 | 83 | 172 | 208 | 83 | 172 | 208 | 83 | 170 | 205 | 83 |
| Sub-Saharan Africa | 224 | 277 | 81 | 225 | 278 | 81 | 228 | 278 | 82 | 229 | 277 | 82 | 227 | 278 | 82 | 227 | 278 | 82 | 225 | 274 | 82 |
| Missing | 309 | 356 | 87 | 304 | 352 | 87 | 297 | 343 | 87 | 290 | 334 | 87 | 281 | 329 | 86 | 274 | 322 | 85 | 266 | 312 | 85 |
| English language proficiency |  |  |  |  |  |  |  |  |  |  |  |  |  |  |  |  |  |  |  |  |  |
| Proficient | 12,193 | 14,601 | 84 | 12,241 | 14,579 | 84 | 12,259 | 14,478 | 85 | 12,195 | 14,358 | 85 | 12,072 | 14,350 | 84 | 11,974 | 14,257 | 84 | 11,800 | 14,006 | 84 |
| Speaks English not well or not at all | 438 | 481 | 91 | 437 | 480 | 91 | 432 | 473 | 91 | 424 | 465 | 91 | 415 | 458 | 91 | 410 | 452 | 91 | 400 | 442 | 91 |
| Missing | 125 | 140 | 89 | 120 | 136 | 88 | 113 | 128 | 89 | 108 | 122 | 89 | 102 | 117 | 88 | 98 | 112 | 87 | 93 | 107 | 87 |
| SEIFA IRSD |  |  |  |  |  |  |  |  |  |  |  |  |  |  |  |  |  |  |  |  |  |
| Most disadvantaged | 3,205 | 3,866 | 83 | 3,178 | 3,851 | 83 | 3,156 | 3,817 | 83 | 3,109 | 3,777 | 82 | 3,026 | 3,766 | 80 | 2,984 | 3,738 | 80 | 2,936 | 3,661 | 80 |
| Quintile 2 | 3,149 | 3,884 | 81 | 3,138 | 3,874 | 81 | 3,128 | 3,845 | 81 | 3,091 | 3,810 | 81 | 3,017 | 3,808 | 79 | 2,981 | 3,785 | 79 | 2,937 | 3,710 | 79 |
| Quintile 3 | 3,207 | 3,937 | 81 | 3,201 | 3,929 | 81 | 3,200 | 3,906 | 82 | 3,166 | 3,874 | 82 | 3,088 | 3,872 | 80 | 3,057 | 3,853 | 79 | 3,019 | 3,787 | 80 |
| Quintile 4 | 3,215 | 3,989 | 81 | 3,216 | 3,984 | 81 | 3,226 | 3,964 | 81 | 3,198 | 3,935 | 81 | 3,121 | 3,935 | 79 | 3,104 | 3,923 | 79 | 3,077 | 3,864 | 80 |
| Least disadvantaged | 3,145 | 4,004 | 79 | 3,151 | 3,998 | 79 | 3,173 | 3,977 | 80 | 3,157 | 3,950 | 80 | 3,088 | 3,955 | 78 | 3,085 | 3,944 | 78 | 3,077 | 3,885 | 79 |
| Missing | 23 | 29 | 80 | 23 | 29 | 80 | 23 | 28 | 81 | 23 | 28 | 81 | 23 | 28 | 80 | 22 | 28 | 80 | 22 | 27 | 80 |
| Household income |  |  |  |  |  |  |  |  |  |  |  |  |  |  |  |  |  |  |  |  |  |
| $104K+ | 1,399 | 1,829 | 76 | 1,408 | 1,828 | 77 | 1,420 | 1,822 | 78 | 1,418 | 1,813 | 78 | 1,392 | 1,819 | 77 | 1,396 | 1,818 | 77 | 1,404 | 1,799 | 78 |
| $65K-<$104K | 2,916 | 3,695 | 79 | 2,927 | 3,696 | 79 | 2,952 | 3,690 | 80 | 2,940 | 3,676 | 80 | 2,878 | 3,690 | 78 | 2,873 | 3,690 | 78 | 2,867 | 3,646 | 79 |
| $26K-$65K | 6,556 | 8,104 | 81 | 6,551 | 8,101 | 81 | 6,567 | 8,072 | 81 | 6,508 | 8,023 | 81 | 6,351 | 8,036 | 79 | 6,300 | 8,009 | 79 | 6,229 | 7,873 | 79 |
| $1-<$26K | 2,944 | 3,461 | 85 | 2,914 | 3,448 | 85 | 2,886 | 3,412 | 85 | 2,835 | 3,366 | 84 | 2,752 | 3,339 | 82 | 2,704 | 3,301 | 82 | 2,645 | 3,220 | 82 |
| Other | 2,129 | 2,619 | 81 | 2,108 | 2,592 | 81 | 2,080 | 2,543 | 82 | 2,044 | 2,498 | 82 | 1,991 | 2,480 | 80 | 1,961 | 2,454 | 80 | 1,924 | 2,397 | 80 |

Notes: English language proficiency and marital status are measured for people aged 18 years and over at the time of Census 2016. Results for employment status are reported for people aged 15-64 years at the time of Census 2016. Results for highest level of education are reported for people aged 25 years and older.

#### Supplementary Table 3. Proportion of the Census 2016 continuity of care study population with high continuity of care (UPI score of ≥0.7) over the study period and in relation to sociodemographic characteristics.

|  | 2016-17 | 2017-18 | 2018-19 | 2019-20 | 2020-21 | 2021-22 | 2022-23 | 1 Apr 2020 31 Mar 2022 |
| --- | --- | --- | --- | --- | --- | --- | --- | --- |
| Total | 32.5 | 32.5 | 32.3 | 35.0 | 38.7 | 37.3 | 37.2 | 39.1 |
| Sex |  |  |  |  |  |  |  |  |
| Male | 34.7 | 34.6 | 34.4 | 37.1 | 40.9 | 39.6 | 39.7 | 41.3 |
| Female | 30.8 | 30.7 | 30.6 | 33.3 | 36.9 | 35.5 | 35.2 | 37.2 |
| Age Group (years) |  |  |  |  |  |  |  |  |
| 0-14 | 18.6 | 18.5 | 18.9 | 21.9 | 26.9 | 26.1 | 25.7 | 27.8 |
| 15-24 | 18.3 | 18.0 | 18.1 | 20.9 | 24.4 | 23.6 | 24.2 | 24.7 |
| 25-44 | 23.6 | 23.1 | 22.7 | 25.5 | 29.3 | 28.5 | 28.5 | 30.0 |
| 45-69 | 42.2 | 41.3 | 40.4 | 42.5 | 45.2 | 43.1 | 42.7 | 45.4 |
| 70+ | 58.6 | 57.6 | 56.5 | 57.3 | 58.4 | 54.9 | 53.5 | 58.2 |
| Relationship status |  |  |  |  |  |  |  |  |
| Married/de facto | 36.9 | 36.8 | 36.6 | 39.3 | 42.6 | 41.1 | 41.0 | 42.9 |
| Single | 34.0 | 33.6 | 33.3 | 36.0 | 39.3 | 38.1 | 38.3 | 39.7 |
| Missing | 37.5 | 37.7 | 36.6 | 38.3 | 40.7 | 38.9 | 38.6 | 40.9 |
| Employment status |  |  |  |  |  |  |  |  |
| Employed | 27.0 | 27.1 | 27.4 | 30.7 | 34.7 | 34.1 | 34.6 | 35.3 |
| Unemployed | 27.3 | 27.0 | 26.9 | 29.9 | 33.8 | 33.1 | 34.0 | 34.2 |
| Not in the labour force | 36.6 | 36.2 | 35.7 | 38.1 | 41.3 | 39.8 | 39.9 | 41.5 |
| Missing | 31.7 | 31.6 | 31.8 | 34.7 | 38.4 | 37.4 | 38.0 | 38.9 |
| Education Level |  |  |  |  |  |  |  |  |
| University | 31.4 | 31.5 | 31.5 | 34.6 | 38.5 | 37.2 | 36.9 | 39.0 |
| High school and/or other qualification | 35.7 | 35.6 | 35.5 | 38.3 | 41.7 | 40.4 | 40.5 | 42.0 |
| No qualification | 46.1 | 45.8 | 45.4 | 47.5 | 50.1 | 48.2 | 47.8 | 50.2 |
| Missing | 52.3 | 51.7 | 50.8 | 52.0 | 53.9 | 51.6 | 50.7 | 53.9 |
| Remoteness area |  |  |  |  |  |  |  |  |
| Major cities | 32.6 | 32.6 | 32.5 | 35.3 | 39.4 | 38.1 | 37.9 | 39.9 |
| Inner regional | 33.8 | 33.5 | 33.1 | 35.8 | 38.2 | 36.3 | 36.2 | 38.2 |
| Outer regional | 31.6 | 31.4 | 31.3 | 33.3 | 36.0 | 35.1 | 35.2 | 36.3 |
| Remote | 24.0 | 23.2 | 23.3 | 24.9 | 28.1 | 28.2 | 28.5 | 28.6 |
| Very remote | 13.5 | 14.7 | 15.2 | 16.8 | 19.4 | 19.1 | 19.5 | 19.6 |
| Other | 21.1 | 23.7 | 25.5 | 29.3 | 33.8 | 33.5 | 34.7 | 34.5 |
| Country of birth |  |  |  |  |  |  |  |  |
| Aus/NZ | 29.9 | 29.8 | 29.7 | 32.5 | 36.1 | 34.7 | 34.6 | 36.4 |
| Other Oceania | 34.4 | 34.8 | 34.9 | 38.3 | 43.0 | 42.4 | 42.5 | 44.0 |
| North-West Europe | 40.1 | 39.8 | 39.3 | 41.6 | 44.1 | 42.2 | 42.1 | 44.2 |
| Southern and Eastern Europe | 54.0 | 53.9 | 53.2 | 55.0 | 58.2 | 55.9 | 55.3 | 58.2 |
| North Africa and Middle East | 39.3 | 39.4 | 39.5 | 41.9 | 46.0 | 45.4 | 45.8 | 46.6 |
| South-East Asia | 40.6 | 40.8 | 40.9 | 43.5 | 48.0 | 47.6 | 47.8 | 48.9 |
| North East Asia | 38.0 | 38.0 | 38.3 | 41.0 | 45.7 | 45.0 | 45.1 | 46.5 |
| Southern and Central Asia | 33.1 | 32.8 | 32.6 | 36.1 | 41.3 | 39.9 | 40.1 | 42.0 |
| Americas | 32.3 | 32.4 | 32.5 | 35.5 | 39.4 | 37.9 | 37.9 | 39.8 |
| Sub-Saharan Africa | 31.5 | 31.4 | 31.0 | 33.9 | 37.8 | 36.8 | 37.0 | 38.3 |
| Missing | 40.7 | 40.3 | 39.8 | 41.8 | 44.6 | 42.9 | 42.6 | 44.8 |
| English language proficiency |  |  |  |  |  |  |  |  |
| Proficient | 35.2 | 35.0 | 34.8 | 37.4 | 40.8 | 39.3 | 39.3 | 41.1 |
| Speaks English not well or not at all | 52.2 | 52.6 | 52.7 | 54.4 | 57.9 | 57.0 | 57.1 | 58.4 |
| Missing | 46.1 | 45.6 | 44.3 | 45.6 | 47.8 | 46.0 | 45.2 | 48.0 |
| SEIFA IRSD |  |  |  |  |  |  |  |  |
| Most disadvantaged | 35.6 | 35.4 | 35.3 | 37.9 | 41.5 | 40.4 | 40.4 | 41.9 |
| Quintile 2 | 33.0 | 32.8 | 32.7 | 35.3 | 38.8 | 37.6 | 37.4 | 39.2 |
| Quintile 3 | 31.8 | 31.9 | 31.8 | 34.6 | 38.3 | 37.0 | 37.1 | 38.7 |
| Quintile 4 | 30.9 | 30.9 | 30.8 | 33.7 | 37.5 | 36.1 | 36.0 | 38.0 |
| Least disadvantaged | 31.4 | 31.3 | 31.0 | 33.6 | 37.4 | 35.7 | 35.2 | 37.8 |
| Missing | 20.7 | 23.4 | 25.2 | 28.9 | 33.5 | 33.3 | 34.4 | 34.2 |
| Household income |  |  |  |  |  |  |  |  |
| $104K+ | 28.0 | 28.2 | 28.3 | 31.3 | 35.4 | 34.2 | 33.8 | 36.0 |
| $65K-<$104K | 27.2 | 27.3 | 27.5 | 30.7 | 34.8 | 33.8 | 33.8 | 35.3 |
| $26K-$65K | 32.0 | 32.0 | 31.9 | 34.7 | 38.5 | 37.2 | 37.2 | 38.9 |
| $1-<$26K | 39.6 | 39.3 | 39.0 | 41.3 | 44.4 | 42.6 | 42.4 | 44.6 |
| Other | 34.7 | 34.5 | 33.9 | 36.1 | 39.1 | 37.6 | 37.4 | 39.4 |
| Frequency of GP visits |  |  |  |  |  |  |  |  |
| 4-6 visits | 30.6 | 30.3 | 30.0 | 32.0 | 35.5 | 34.8 | 35.0 | 36.2 |
| 7-9 visits | 30.0 | 29.8 | 29.6 | 32.2 | 35.9 | 34.8 | 34.8 | 36.4 |
| 10-14 visits | 30.7 | 30.6 | 30.5 | 33.3 | 37.0 | 35.7 | 35.5 | 37.5 |
| 15-19 visits | 31.9 | 32.0 | 31.9 | 34.7 | 38.3 | 36.6 | 36.4 | 38.5 |
| 20-39 visits | 37.6 | 37.6 | 37.5 | 40.1 | 43.2 | 41.4 | 41.3 | 43.3 |
| 40+ visits | 43.8 | 43.2 | 42.8 | 45.7 | 48.9 | 47.2 | 47.4 | 48.9 |

Notes: English language proficiency and marital status are measured for people aged 18 years and over at the time of Census 2021. Results for employment status are reported for people aged 15-64 years at the time of Census 2021. Results for highest level of education are reported for people aged 25 years and older.

#### Supplementary Table 4. Comparison of proportions of the total MBS and Census 2016 study population with at least 4 GP visits (UPI coverage), 2016-17 to 2022-23.

|  | 2016-17 | | | 2017-18 | | | 2018-19 | | | 2019-20 | | | 2020-21 | | | 2021-22 | | | 2022-23 | | |
| --- | --- | --- | --- | --- | --- | --- | --- | --- | --- | --- | --- | --- | --- | --- | --- | --- | --- | --- | --- | --- | --- |
|  | MBS | Cen. | Δ | MBS | Cen. | Δ | MBS | Cen. | Δ | MBS | Cen. | Δ | MBS | Cen. | Δ | MBS | Cen. | Δ | MBS | Cen. | Δ |
| Total | 79.4 | 80.9 | 1.5 | 79.8 | 80.9 | 1.1 | 80.4 | 81.4 | 1.0 | 80.4 | 81.3 | 0.9 | 78.4 | 79.3 | 0.9 | 78.4 | 79.0 | 0.7 | 78.7 | 79.6 | 0.9 |
| Sex |  |  |  |  |  |  |  |  |  |  |  |  |  |  |  |  |  |  |  |  |  |
| Male | 74.5 | 75.8 | 1.3 | 74.9 | 75.8 | 0.9 | 75.7 | 76.4 | 0.7 | 75.7 | 76.2 | 0.5 | 73.2 | 73.7 | 0.5 | 73.0 | 73.2 | 0.2 | 73.3 | 73.7 | 0.4 |
| Female | 84.2 | 85.7 | 1.5 | 84.5 | 85.7 | 1.2 | 85.0 | 86.1 | 1.1 | 84.9 | 86.0 | 1.1 | 83.4 | 84.6 | 1.2 | 83.6 | 84.5 | 1.0 | 83.8 | 85.0 | 1.2 |
| Age Group (years) | |  |  |  |  |  |  |  |  |  |  |  |  |  |  |  |  |  |  |  |  |
| 0-14 | 70.3 | 71.2 | 0.8 | 70.3 | 69.2 | -1.1 | 71.1 | 68.6 | -2.5 | 69.9 | 66.2 | -3.7 | 64.3 | 58.3 | -6.0 | 64.9 | 56.2 | -8.7 | 65.1 | 57.7 | -7.4 |
| 15-24 | 73.7 | 74.9 | 1.2 | 73.9 | 74.9 | 1.0 | 74.5 | 75.4 | 1.0 | 74.1 | 74.8 | 0.7 | 71.5 | 72.1 | 0.6 | 70.6 | 71.2 | 0.5 | 70.2 | 70.7 | 0.5 |
| 25-44 | 77.1 | 78.8 | 1.7 | 77.6 | 79.2 | 1.5 | 78.4 | 79.9 | 1.5 | 78.6 | 79.8 | 1.2 | 76.9 | 78.0 | 1.1 | 76.3 | 77.2 | 0.9 | 76.3 | 77.1 | 0.7 |
| 45-69 | 85.3 | 86.3 | 1.0 | 85.6 | 86.5 | 0.8 | 86.0 | 86.8 | 0.8 | 86.2 | 87.0 | 0.8 | 85.7 | 86.4 | 0.7 | 85.5 | 86.3 | 0.7 | 85.6 | 86.4 | 0.8 |
| 70+ | 94.6 | 95.8 | 1.1 | 94.8 | 95.3 | 0.5 | 95.0 | 95.4 | 0.5 | 95.1 | 95.5 | 0.4 | 95.2 | 95.5 | 0.3 | 95.3 | 95.6 | 0.3 | 95.2 | 95.6 | 0.3 |
| Remoteness area | |  |  |  |  |  |  |  |  |  |  |  |  |  |  |  |  |  |  |  |  |
| Major cities | 81.0 | 81.9 | 0.9 | 81.3 | 81.8 | 0.5 | 82.1 | 82.4 | 0.4 | 81.9 | 82.2 | 0.3 | 79.8 | 80.1 | 0.3 | 79.5 | 79.9 | 0.4 | 79.8 | 80.4 | 0.6 |
| Inner regional | 78.7 | 79.4 | 0.8 | 79.0 | 79.5 | 0.5 | 79.5 | 79.9 | 0.4 | 79.3 | 79.8 | 0.5 | 77.7 | 78.4 | 0.7 | 77.2 | 78.1 | 0.9 | 77.6 | 78.7 | 1.1 |
| Outer regional | 77.1 | 78.0 | 0.9 | 77.2 | 77.8 | 0.6 | 77.4 | 78.0 | 0.6 | 77.1 | 78.0 | 0.9 | 75.3 | 76.6 | 1.3 | 74.3 | 75.9 | 1.6 | 74.2 | 76.3 | 2.1 |
| Remote | 71.3 | 73.0 | 1.7 | 71.4 | 73.2 | 1.7 | 71.0 | 73.2 | 2.2 | 70.5 | 73.3 | 2.8 | 68.6 | 71.9 | 3.3 | 66.9 | 70.9 | 3.9 | 66.5 | 71.2 | 4.7 |
| Very remote | 67.6 | 66.4 | -1.2 | 68.2 | 67.5 | -0.7 | 68.1 | 68.1 | -0.1 | 67.3 | 68.1 | 0.8 | 65.7 | 67.1 | 1.4 | 64.4 | 66.1 | 1.7 | 64.6 | 66.8 | 2.3 |
| Other | 73.7 | 80.2 | 6.4 | 73.9 | 80.9 | 7.1 | 74.1 | 81.8 | 7.7 | 74.5 | 81.9 | 7.3 | 72.4 | 80.7 | 8.3 | 76.8 | 80.2 | 3.3 | 77.0 | 80.4 | 3.4 |
| SEIFA IRSD |  |  |  |  |  |  |  |  |  |  |  |  |  |  |  |  |  |  |  |  |  |
| Most disadvantaged | 82.6 | 82.9 | 0.3 | 82.5 | 82.5 | 0.1 | 82.7 | 82.7 | 0.0 | 82.2 | 82.3 | 0.1 | 80.0 | 80.3 | 0.3 | 79.3 | 79.8 | 0.5 | 79.4 | 80.2 | 0.8 |
| Quintile 2 | 81.2 | 81.1 | -0.2 | 81.5 | 81.0 | -0.5 | 82.0 | 81.4 | -0.6 | 81.7 | 81.1 | -0.6 | 79.7 | 79.2 | -0.5 | 79.1 | 78.8 | -0.3 | 79.1 | 79.2 | 0.0 |
| Quintile 3 | 80.2 | 81.5 | 1.2 | 80.6 | 81.5 | 0.8 | 81.3 | 81.9 | 0.6 | 81.1 | 81.7 | 0.6 | 79.2 | 79.8 | 0.6 | 78.7 | 79.3 | 0.6 | 78.8 | 79.7 | 0.9 |
| Quintile 4 | 79.1 | 80.6 | 1.5 | 79.6 | 80.7 | 1.1 | 80.5 | 81.4 | 0.9 | 80.4 | 81.3 | 0.9 | 78.5 | 79.3 | 0.8 | 78.3 | 79.1 | 0.9 | 78.5 | 79.6 | 1.1 |
| Least disadvantaged | 77.2 | 78.6 | 1.4 | 77.9 | 78.8 | 1.0 | 78.9 | 79.8 | 0.9 | 79.2 | 79.9 | 0.8 | 77.3 | 78.1 | 0.8 | 77.2 | 78.2 | 1.0 | 78.0 | 79.2 | 1.2 |
| Missing | 73.8 | 79.6 | 5.8 | 74.0 | 80.4 | 6.4 | 74.2 | 81.2 | 7.0 | 74.7 | 81.3 | 6.6 | 72.6 | 80.1 | 7.6 | 76.8 | 79.6 | 2.7 | 77.0 | 79.7 | 2.7 |

Note. Cen. = Census 2016, Δ = difference.

#### Supplementary Table 5. Comparison of proportions of the total MBS continuity of care study population and Census 2016 continuity of care study population with high continuity (UPI score ≥0.7), 2016-17 to 2022-23.

|  | 2016-17 | | | 2017-18 | | | 2018-19 | | | 2019-20 | | | 2020-21 | | | 2021-22 | | | 2022-23 | | |
| --- | --- | --- | --- | --- | --- | --- | --- | --- | --- | --- | --- | --- | --- | --- | --- | --- | --- | --- | --- | --- | --- |
|  | MBS | Cen. | Δ | MBS | Cen. | Δ | MBS | Cen. | Δ | MBS | Cen. | Δ | MBS | Cen. | Δ | MBS | Cen. | Δ | MBS | Cen. | Δ |
| Total | 32.1 | 32.5 | 0.5 | 31.7 | 32.5 | 0.8 | 31.3 | 32.3 | 1.1 | 33.8 | 35.0 | 1.2 | 37.2 | 38.7 | 1.5 | 35.7 | 37.3 | 1.6 | 35.5 | 37.2 | 1.7 |
| Sex |  |  |  |  |  |  |  |  |  |  |  |  |  |  |  |  |  |  |  |  |  |
| Male | 34.1 | 34.7 | 0.6 | 33.6 | 34.6 | 1.0 | 33.1 | 34.4 | 1.3 | 35.6 | 37.1 | 1.5 | 39.0 | 40.9 | 1.8 | 37.6 | 39.6 | 2.0 | 37.6 | 39.7 | 2.1 |
| Female | 30.4 | 30.8 | 0.4 | 30.0 | 30.7 | 0.7 | 29.7 | 30.6 | 0.9 | 32.3 | 33.3 | 1.0 | 35.7 | 36.9 | 1.2 | 34.1 | 35.5 | 1.4 | 33.8 | 35.2 | 1.4 |
| Age Group (years) | |  |  |  |  |  |  |  |  |  |  |  |  |  |  |  |  |  |  |  |  |
| 0-14 | 19.2 | 18.6 | -0.6 | 19.0 | 18.5 | -0.5 | 18.9 | 18.9 | 0.0 | 21.9 | 21.9 | 0.0 | 25.6 | 26.9 | 1.4 | 23.5 | 26.1 | 2.6 | 22.4 | 25.7 | 3.3 |
| 15-24 | 17.9 | 18.3 | 0.4 | 17.7 | 18.0 | 0.3 | 17.8 | 18.1 | 0.2 | 20.7 | 20.9 | 0.2 | 24.2 | 24.4 | 0.2 | 23.6 | 23.6 | 0.0 | 24.1 | 24.2 | 0.0 |
| 25-44 | 23.5 | 23.6 | 0.2 | 23.0 | 23.1 | 0.1 | 22.6 | 22.7 | 0.1 | 25.5 | 25.5 | 0.0 | 29.2 | 29.3 | 0.0 | 28.5 | 28.5 | 0.0 | 28.5 | 28.5 | -0.1 |
| 45-69 | 42.0 | 42.2 | 0.2 | 41.1 | 41.3 | 0.2 | 40.3 | 40.4 | 0.1 | 42.4 | 42.5 | 0.1 | 45.1 | 45.2 | 0.1 | 43.1 | 43.1 | 0.1 | 42.7 | 42.7 | 0.0 |
| 70+ | 58.5 | 58.6 | 0.0 | 57.5 | 57.6 | 0.1 | 56.3 | 56.5 | 0.1 | 57.1 | 57.3 | 0.1 | 58.3 | 58.4 | 0.1 | 54.8 | 54.9 | 0.1 | 53.5 | 53.5 | 0.0 |
| Remoteness area |  |  |  |  |  |  |  |  |  |  |  |  |  |  |  |  |  |  |  |  |  |
| Major cities | 32.4 | 32.6 | 0.2 | 32.0 | 32.6 | 0.6 | 31.6 | 32.5 | 0.9 | 34.1 | 35.3 | 1.2 | 38.0 | 39.4 | 1.4 | 36.6 | 38.1 | 1.5 | 36.3 | 37.9 | 1.6 |
| Inner regional | 33.2 | 33.8 | 0.7 | 32.5 | 33.5 | 1.0 | 32.1 | 33.1 | 1.1 | 34.5 | 35.8 | 1.3 | 36.8 | 38.2 | 1.4 | 34.8 | 36.3 | 1.5 | 34.6 | 36.2 | 1.7 |
| Outer regional | 29.7 | 31.6 | 1.9 | 29.3 | 31.4 | 2.1 | 29.0 | 31.3 | 2.3 | 31.1 | 33.3 | 2.2 | 33.8 | 36.0 | 2.2 | 32.9 | 35.1 | 2.2 | 33.1 | 35.2 | 2.1 |
| Remote | 23.5 | 24.0 | 0.5 | 22.6 | 23.2 | 0.6 | 22.3 | 23.3 | 1.0 | 23.8 | 24.9 | 1.1 | 26.7 | 28.1 | 1.4 | 26.6 | 28.2 | 1.6 | 26.9 | 28.5 | 1.6 |
| Very remote | 14.1 | 13.5 | -0.5 | 14.4 | 14.7 | 0.3 | 14.1 | 15.2 | 1.1 | 15.2 | 16.8 | 1.6 | 16.7 | 19.4 | 2.6 | 16.2 | 19.1 | 2.9 | 15.3 | 19.5 | 4.3 |
| Other | 31.2 | 21.1 | -10.1 | 30.8 | 23.7 | -7.1 | 30.5 | 25.5 | -5.1 | 34.2 | 29.3 | -4.9 | 36.1 | 33.8 | -2.3 | 33.4 | 33.5 | 0.2 | 33.9 | 34.7 | 0.8 |
| SEIFA IRSD |  |  |  |  |  |  |  |  |  |  |  |  |  |  |  |  |  |  |  |  |  |
| Most disadvantaged | 34.4 | 35.6 | 1.2 | 34.0 | 35.4 | 1.3 | 33.9 | 35.3 | 1.4 | 36.4 | 37.9 | 1.6 | 40.1 | 41.5 | 1.4 | 39.0 | 40.4 | 1.3 | 39.1 | 40.4 | 1.3 |
| Quintile 2 | 32.4 | 33.0 | 0.6 | 32.0 | 32.8 | 0.8 | 31.6 | 32.7 | 1.1 | 34.2 | 35.3 | 1.1 | 37.7 | 38.8 | 1.1 | 36.4 | 37.6 | 1.1 | 36.4 | 37.4 | 1.0 |
| Quintile 3 | 31.5 | 31.8 | 0.4 | 31.1 | 31.9 | 0.9 | 30.7 | 31.8 | 1.2 | 33.2 | 34.6 | 1.4 | 36.7 | 38.3 | 1.6 | 35.2 | 37.0 | 1.8 | 35.1 | 37.1 | 2.0 |
| Quintile 4 | 31.2 | 30.9 | -0.3 | 30.7 | 30.9 | 0.3 | 30.1 | 30.8 | 0.7 | 32.6 | 33.7 | 1.0 | 36.1 | 37.5 | 1.5 | 34.5 | 36.1 | 1.7 | 34.1 | 36.0 | 1.9 |
| Least disadvantaged | 31.1 | 31.4 | 0.3 | 30.6 | 31.3 | 0.7 | 30.0 | 31.0 | 1.0 | 32.4 | 33.6 | 1.3 | 35.8 | 37.4 | 1.5 | 34.1 | 35.7 | 1.6 | 33.4 | 35.2 | 1.8 |
| Missing | 31.3 | 20.7 | -10.5 | 30.9 | 23.4 | -7.5 | 30.7 | 25.2 | -5.5 | 34.3 | 28.9 | -5.4 | 36.2 | 33.5 | -2.7 | 33.5 | 33.3 | -0.2 | 34.2 | 34.4 | 0.2 |

Note. Cen. = Census 2016, Δ = difference. The MBS study population includes all individuals with at least 4 claims for GP services in each of the 2-year study periods. The Census 2016 study population includes individuals with a 2016 Census record that linked to the PLIDA Spine who had at least 4 claims for GP services in each of the 2-year study period.

#### Supplementary Table 6. Proportions of the Census 2021 continuity of care study population with high continuity when using different definitions of high, 2022-23.

|  | Cut point | | |
| --- | --- | --- | --- |
|  | ≥0.70 | ≥0.75 | ≥0.80 |
| Total | 36.0 | 31.5 | 25.0 |
| Sex |  |  |  |
| Male | 38.2 | 33.9 | 27.1 |
| Female | 34.2 | 29.5 | 23.3 |
| Age Group (years) |  |  |  |
| 0-14 | 22.7 | 19.9 | 14.3 |
| 15-24 | 24.4 | 21.2 | 15.7 |
| 25-44 | 28.7 | 24.8 | 18.8 |
| 45-69 | 42.7 | 37.6 | 30.5 |
| 70+ | 53.4 | 47.1 | 39.3 |
| Relationship status |  |  |  |
| Married/de facto | 39.2 | 34.2 | 27.4 |
| Single | 37.6 | 33.0 | 26.6 |
| Missing | 40.6 | 35.9 | 29.6 |
| Employment status |  |  |  |
| Employed | 31.8 | 27.6 | 21.4 |
| Unemployed | 32.8 | 28.8 | 22.7 |
| Not in the labour force | 40.2 | 35.5 | 28.8 |
| Missing | 35.6 | 31.4 | 25.2 |
| Education Level |  |  |  |
| University | 35.1 | 30.5 | 23.9 |
| High school and/or other qualification | 39.1 | 34.2 | 27.4 |
| No qualification | 47.7 | 42.1 | 34.9 |
| Missing | 51.8 | 46.4 | 39.5 |
| Remoteness area |  |  |  |
| Major cities | 36.6 | 32.1 | 25.6 |
| Inner regional | 34.9 | 30.3 | 23.7 |
| Outer regional | 33.6 | 29.4 | 23.1 |
| Remote | 27.7 | 24.2 | 18.3 |
| Very remote | 15.7 | 13.4 | 9.5 |
| Other | 36.3 | 31.7 | 25.1 |
| Country of birth |  |  |  |
| Aus/NZ | 33.1 | 28.8 | 22.5 |
| Other Oceania | 42.1 | 37.3 | 30.6 |
| North-West Europe | 41.1 | 35.8 | 28.7 |
| Southern and Eastern Europe | 54.7 | 49.3 | 42.2 |
| North Africa and Middle East | 46.0 | 40.9 | 34.1 |
| South-East Asia | 46.9 | 42.2 | 35.3 |
| North East Asia | 44.3 | 39.8 | 33.1 |
| Southern and Central Asia | 39.1 | 34.3 | 27.8 |
| Americas | 36.4 | 31.7 | 25.2 |
| Sub-Saharan Africa | 36.6 | 31.9 | 25.2 |
| Missing | 44.6 | 39.5 | 32.6 |
| English language proficiency |  |  |  |
| Proficient | 38.0 | 33.2 | 26.5 |
| Speaks English not well or not at all | 57.5 | 52.2 | 45.2 |
| Missing | 48.3 | 43.0 | 36.4 |
| SEIFA IRSD |  |  |  |
| Most disadvantaged | 39.9 | 35.3 | 28.8 |
| Quintile 2 | 36.9 | 32.4 | 25.9 |
| Quintile 3 | 35.4 | 30.9 | 24.5 |
| Quintile 4 | 34.4 | 30.0 | 23.5 |
| Least disadvantaged | 33.5 | 29.1 | 22.7 |
| Missing | 36.5 | 31.9 | 25.2 |
| Household income |  |  |  |
| $104K+ | 31.2 | 27.1 | 20.8 |
| $65K-<$104K | 31.8 | 27.6 | 21.4 |
| $26K-$65K | 37.1 | 32.5 | 25.9 |
| $1-<$26K | 43.7 | 38.6 | 31.7 |
| Other | 38.6 | 34.1 | 27.6 |
| Health conditions |  |  |  |
| Arthritis | 48.6 | 42.5 | 35.1 |
| Asthma | 35.3 | 30.6 | 24.3 |
| Cancer | 48.8 | 42.7 | 35.1 |
| Dementia | 59.0 | 53.4 | 46.5 |
| Diabetes | 53.6 | 47.6 | 40.2 |
| Heart disease | 52.3 | 46.1 | 38.6 |
| Kidney disease | 51.3 | 45.3 | 38.0 |
| Lung disease | 50.7 | 44.5 | 36.9 |
| Mental health condition | 38.0 | 32.9 | 26.6 |
| Stroke | 53.2 | 47.1 | 39.7 |
| Number of named conditions |  |  |  |
| None of the conditions | 32.3 | 28.3 | 22.0 |
| 1 condition | 40.0 | 34.9 | 28.2 |
| 2 conditions | 46.4 | 40.6 | 33.5 |
| 3 or more conditions | 52.7 | 46.5 | 38.9 |
| Frequency of GP visits |  |  |  |
| 4-6 visits | 33.9 | 33.9 | 25.4 |
| 7-9 visits | 33.6 | 28.3 | 20.7 |
| 10-14 visits | 34.3 | 28.2 | 23.7 |
| 15-19 visits | 35.2 | 29.9 | 24.8 |
| 20-39 visits | 40.2 | 34.4 | 28.1 |
| 40+ visits | 46.6 | 40.4 | 33.5 |

Notes: English language proficiency and marital status are measured for people aged 18 years and over at the time of Census 2021. Results for employment status are reported for people aged 15-64 years at the time of Census 2021. Results for highest level of education are reported for people aged 25 years and older.

#### Supplementary Table 7. Descriptive statistics for continuity of care, measured with continuous Usual Provider Index scores, for the Census 2021 continuity of care study population, 2022-23.

|  | Mean | Median | 25^th^ percentile | 75^th^ percentile |
| --- | --- | --- | --- | --- |
| Total | 0.60 | 0.57 | 0.40 | 0.80 |
| Sex |  |  |  |  |
| Male | 0.61 | 0.60 | 0.42 | 0.80 |
| Female | 0.59 | 0.57 | 0.40 | 0.78 |
| Age Group (years) |  |  |  |  |
| 0-14 | 0.51 | 0.50 | 0.33 | 0.67 |
| 15-24 | 0.53 | 0.50 | 0.33 | 0.68 |
| 25-44 | 0.56 | 0.50 | 0.38 | 0.74 |
| 45-69 | 0.64 | 0.63 | 0.45 | 0.83 |
| 70+ | 0.70 | 0.72 | 0.53 | 0.88 |
| Relationship status |  |  |  |  |
| Married/de facto | 0.62 | 0.60 | 0.43 | 0.80 |
| Single | 0.61 | 0.59 | 0.41 | 0.80 |
| Missing | 0.62 | 0.61 | 0.43 | 0.83 |
| Employment status |  |  |  |  |
| Employed | 0.58 | 0.55 | 0.40 | 0.75 |
| Unemployed | 0.58 | 0.55 | 0.40 | 0.76 |
| Not in the labour force | 0.62 | 0.60 | 0.43 | 0.82 |
| Missing | 0.59 | 0.57 | 0.40 | 0.80 |
| Education Level |  |  |  |  |
| University | 0.60 | 0.57 | 0.41 | 0.78 |
| High school and/or other qualification | 0.62 | 0.60 | 0.43 | 0.81 |
| No qualification | 0.67 | 0.67 | 0.50 | 0.86 |
| Missing | 0.69 | 0.71 | 0.50 | 0.89 |
| Remoteness area |  |  |  |  |
| Major cities | 0.60 | 0.58 | 0.40 | 0.80 |
| Inner regional | 0.59 | 0.57 | 0.40 | 0.78 |
| Outer regional | 0.59 | 0.56 | 0.40 | 0.77 |
| Remote | 0.55 | 0.50 | 0.36 | 0.73 |
| Very remote | 0.45 | 0.40 | 0.29 | 0.58 |
| Other | 0.60 | 0.58 | 0.40 | 0.80 |
| Country of birth |  |  |  |  |
| Aus/NZ | 0.58 | 0.56 | 0.40 | 0.76 |
| Other Oceania | 0.63 | 0.63 | 0.44 | 0.83 |
| North-West Europe | 0.63 | 0.63 | 0.44 | 0.82 |
| Southern and Eastern Europe | 0.70 | 0.74 | 0.51 | 0.90 |
| North Africa and Middle East | 0.66 | 0.67 | 0.47 | 0.86 |
| South-East Asia | 0.67 | 0.67 | 0.50 | 0.88 |
| North East Asia | 0.65 | 0.64 | 0.46 | 0.86 |
| Southern and Central Asia | 0.62 | 0.60 | 0.43 | 0.81 |
| Americas | 0.60 | 0.58 | 0.41 | 0.80 |
| Sub-Saharan Africa | 0.60 | 0.59 | 0.42 | 0.80 |
| Missing | 0.65 | 0.65 | 0.45 | 0.85 |
| English language proficiency |  |  |  |  |
| Proficient | 0.61 | 0.60 | 0.42 | 0.80 |
| Speaks English not well or not at all | 0.73 | 0.75 | 0.55 | 0.93 |
| Missing | 0.67 | 0.68 | 0.48 | 0.88 |
| SEIFA IRSD |  |  |  |  |
| Most disadvantaged | 0.62 | 0.60 | 0.43 | 0.82 |
| Quintile 2 | 0.60 | 0.58 | 0.40 | 0.80 |
| Quintile 3 | 0.59 | 0.57 | 0.40 | 0.79 |
| Quintile 4 | 0.59 | 0.57 | 0.40 | 0.78 |
| Least disadvantaged | 0.58 | 0.56 | 0.40 | 0.76 |
| Missing | 0.60 | 0.58 | 0.40 | 0.80 |
| Household income |  |  |  |  |
| $104K+ | 0.57 | 0.54 | 0.40 | 0.75 |
| $65K-<$104K | 0.57 | 0.55 | 0.40 | 0.75 |
| $26K-$65K | 0.60 | 0.59 | 0.41 | 0.80 |
| $1-<$26K | 0.64 | 0.64 | 0.45 | 0.84 |
| Other | 0.61 | 0.60 | 0.42 | 0.81 |
| Health conditions |  |  |  |  |
| Arthritis | 0.67 | 0.69 | 0.50 | 0.86 |
| Asthma | 0.59 | 0.57 | 0.40 | 0.79 |
| Cancer | 0.67 | 0.69 | 0.50 | 0.86 |
| Dementia | 0.73 | 0.77 | 0.56 | 0.92 |
| Diabetes | 0.70 | 0.73 | 0.52 | 0.89 |
| Heart disease | 0.69 | 0.71 | 0.50 | 0.88 |
| Kidney disease | 0.68 | 0.71 | 0.50 | 0.88 |
| Lung disease | 0.68 | 0.70 | 0.50 | 0.87 |
| Mental health condition | 0.61 | 0.60 | 0.42 | 0.80 |
| Stroke | 0.70 | 0.72 | 0.52 | 0.89 |
| Number of named conditions |  |  |  |  |
| None of the conditions | 0.58 | 0.55 | 0.40 | 0.75 |
| 1 condition | 0.62 | 0.61 | 0.43 | 0.82 |
| 2 conditions | 0.66 | 0.67 | 0.47 | 0.86 |
| 3 or more conditions | 0.69 | 0.72 | 0.52 | 0.88 |
| Frequency of GP visits |  |  |  |  |
| 4-6 visits | 0.60 | 0.50 | 0.40 | 0.80 |
| 7-9 visits | 0.58 | 0.56 | 0.38 | 0.75 |
| 10-14 visits | 0.58 | 0.55 | 0.40 | 0.77 |
| 15-19 visits | 0.59 | 0.58 | 0.40 | 0.79 |
| 20-39 visits | 0.62 | 0.62 | 0.43 | 0.82 |
| 40+ visits | 0.66 | 0.67 | 0.48 | 0.86 |

Notes: English language proficiency and marital status are measured for people aged 18 years and over at the time of Census 2021. Results for employment status are reported for people aged 15-64 years at the time of Census 2021. Results for highest level of education are reported for people aged 25 years and older.

### Supplementary Figures

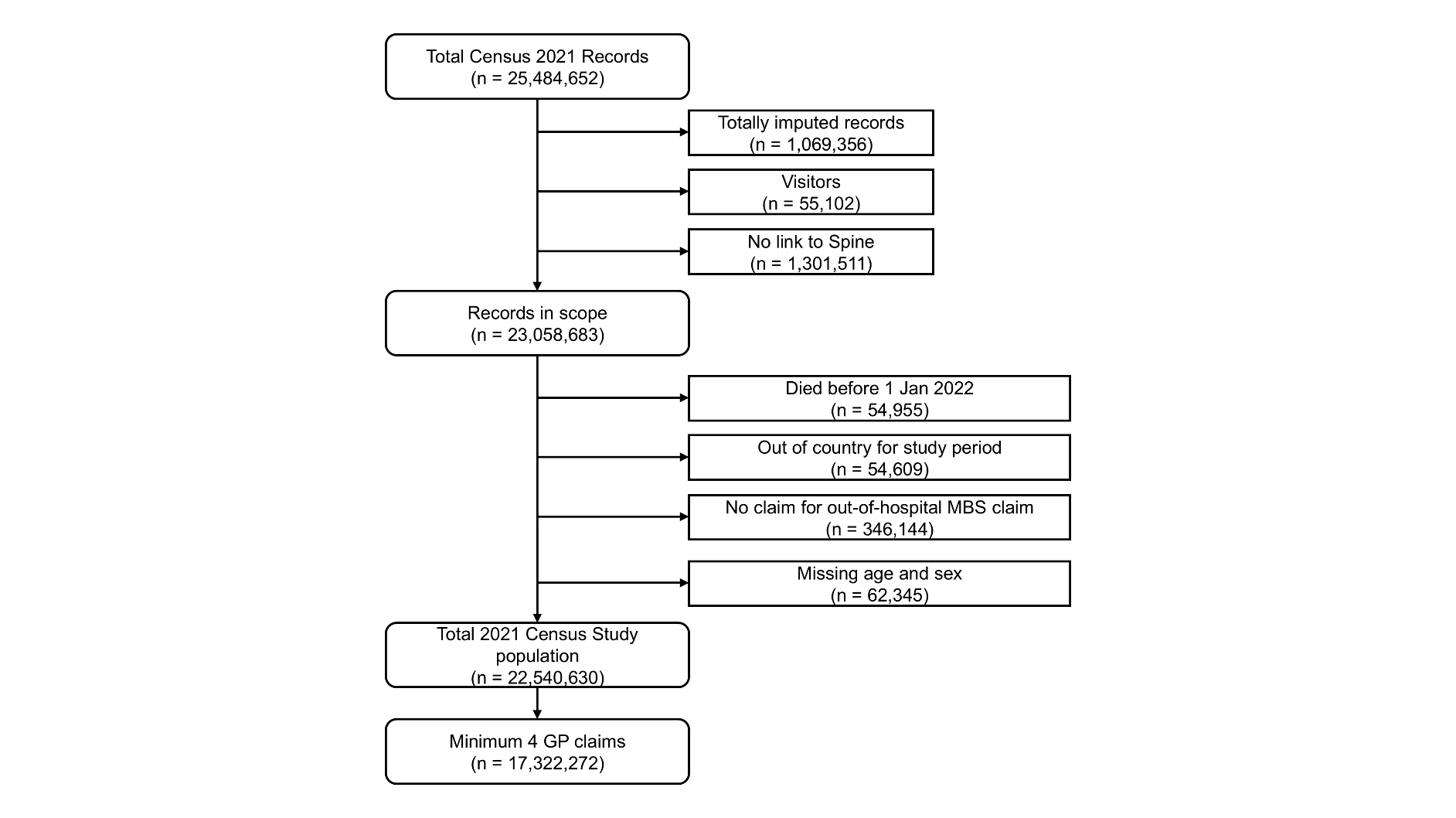

Supplementary Figure 1. Study flow diagram for the study population for the 2022-23 study period, based on the 2021 Census.

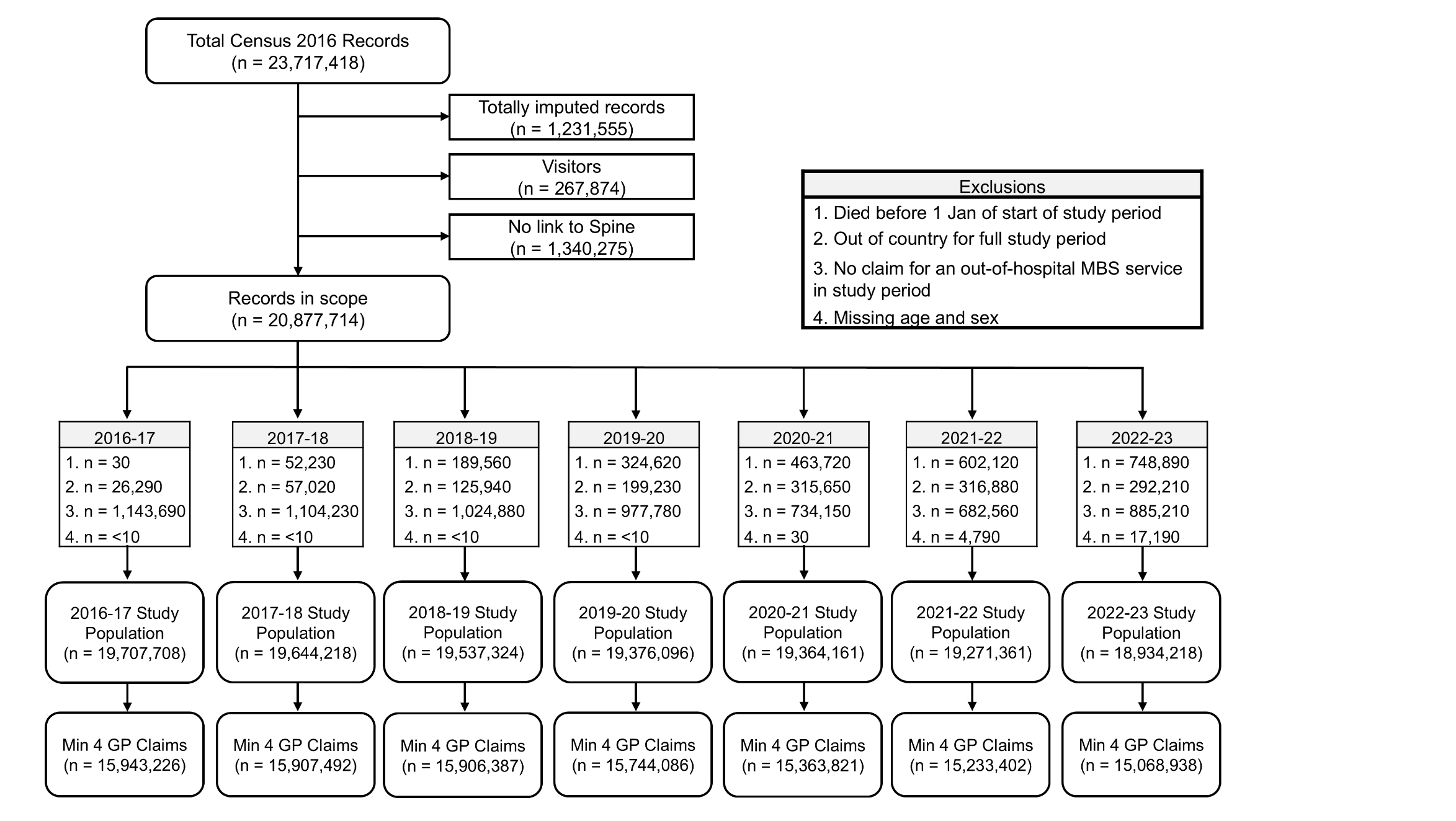

#### Supplementary Figure 2. Study flow diagram for the study populations for each of the study periods, from 2016-17 to 2022-23, based on Census 2016

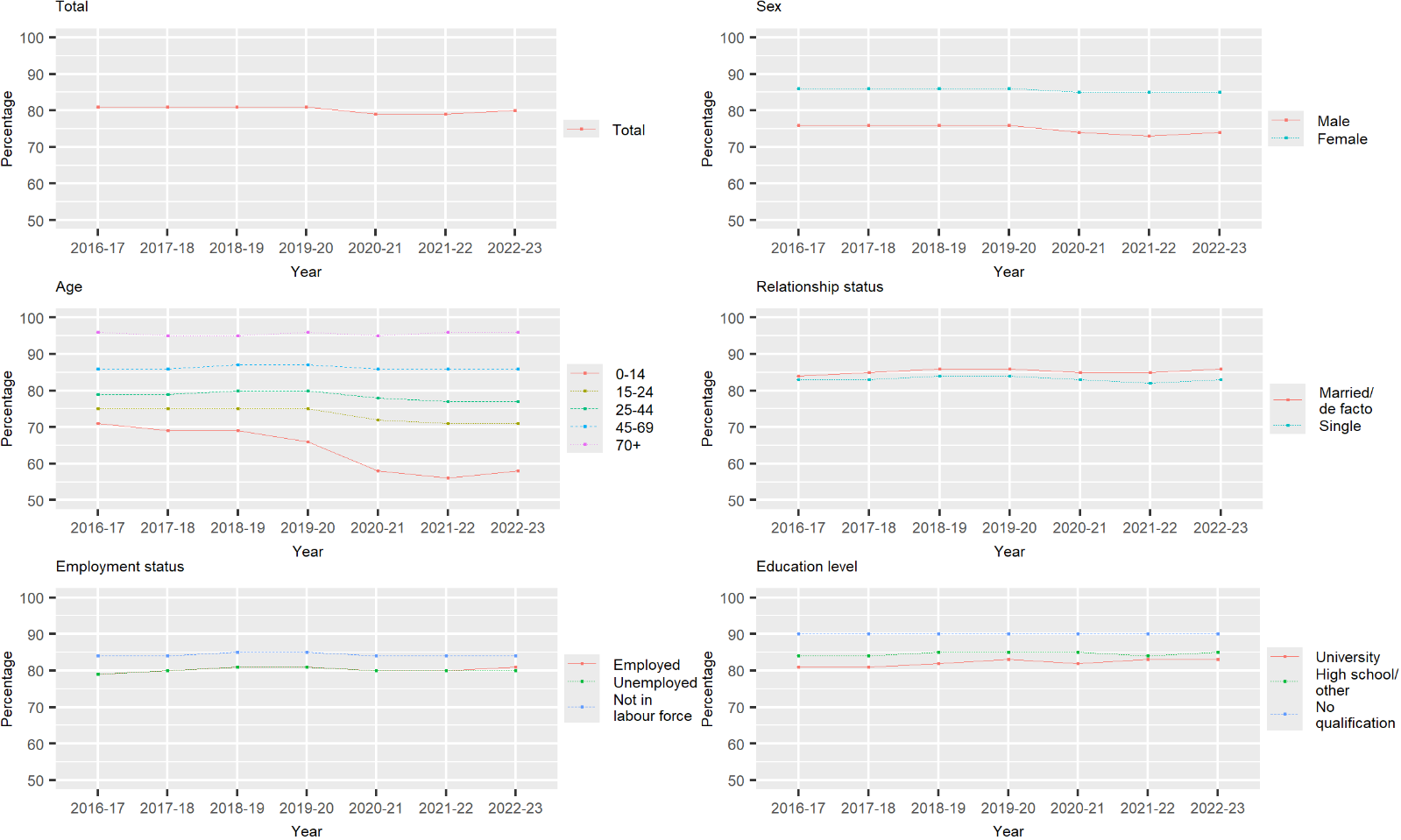

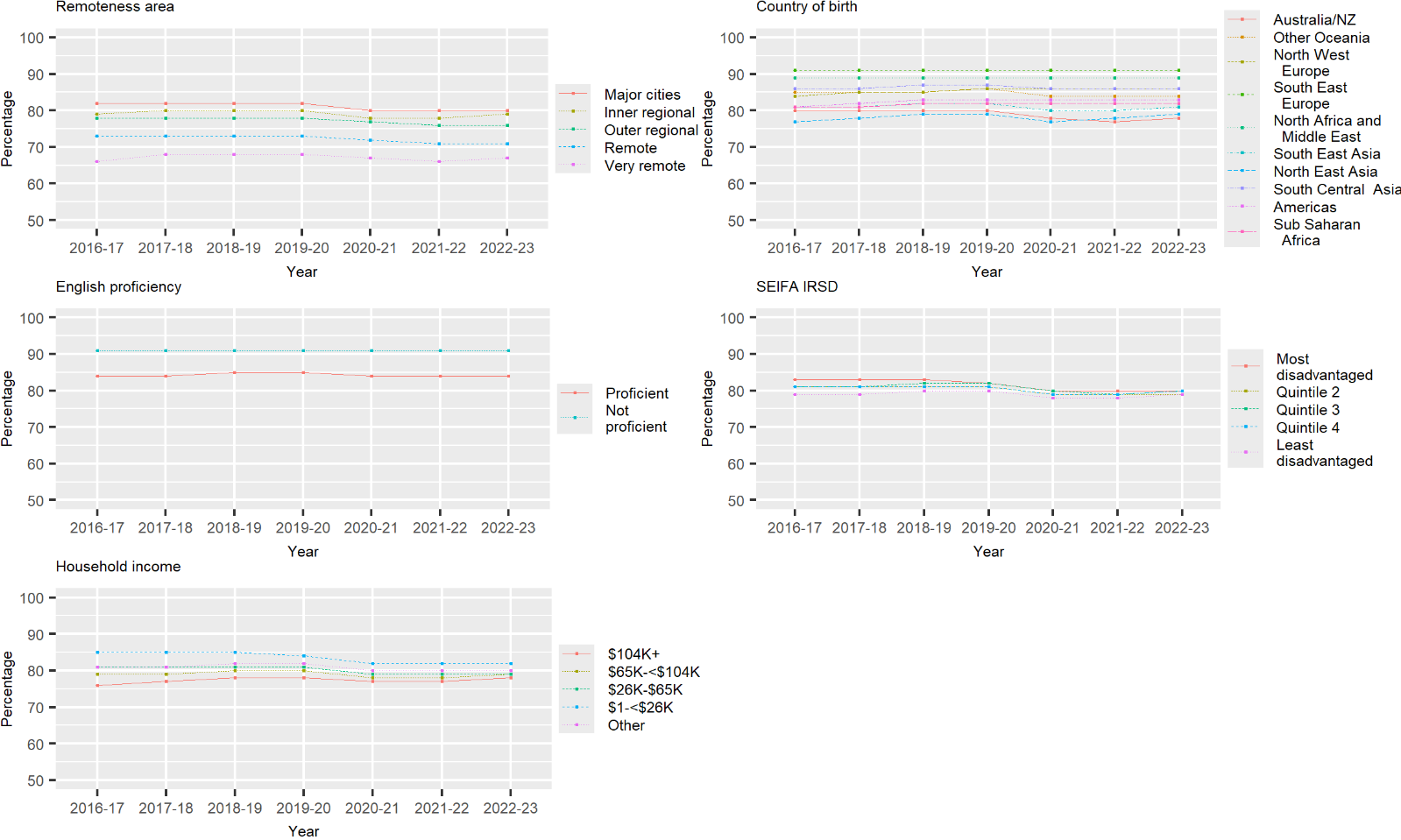

#### Supplementary Figure 3. Proportions of the Census 2016 study population with at least 4 GP visits in each of the 2-year study periods, by sociodemographic characteristics

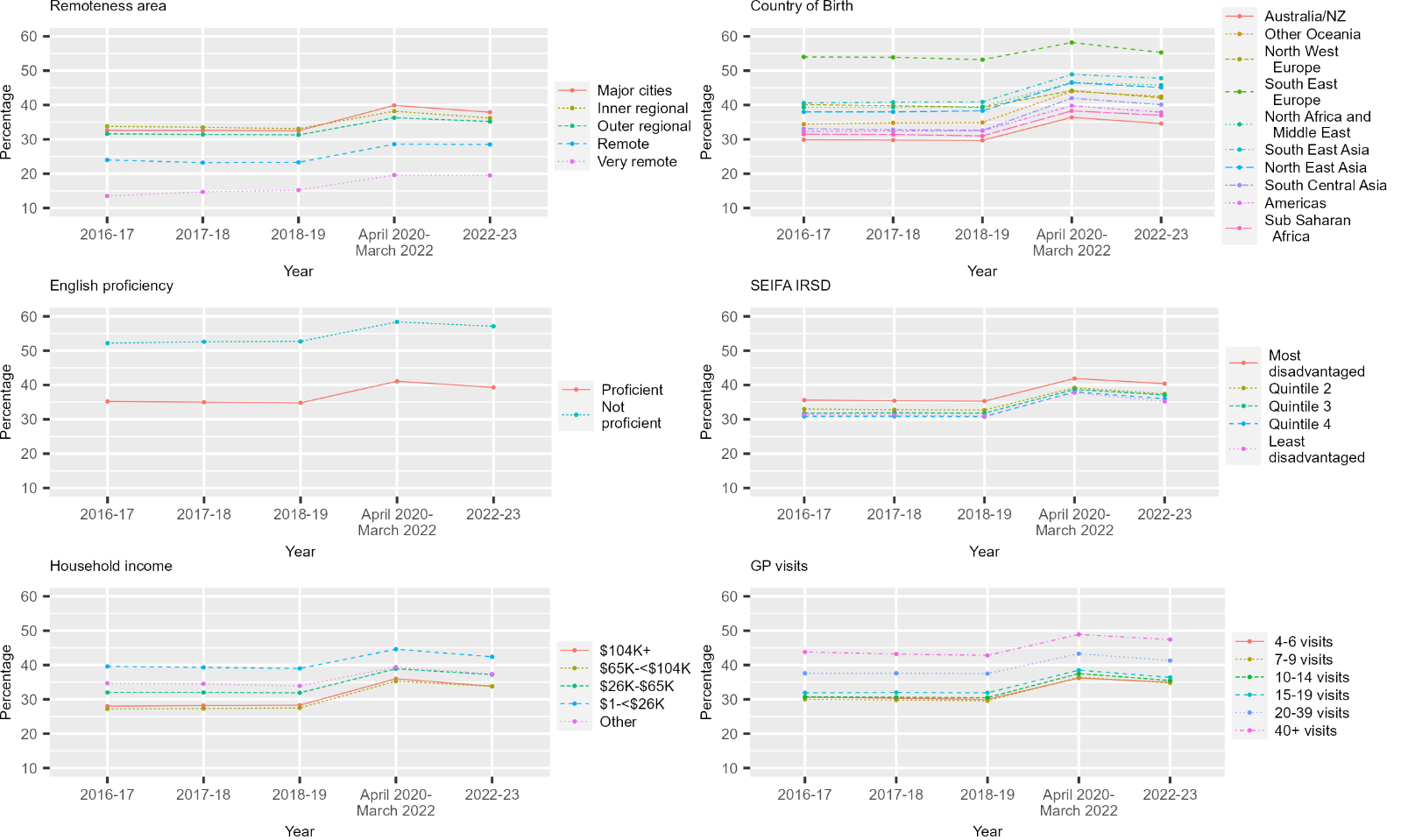

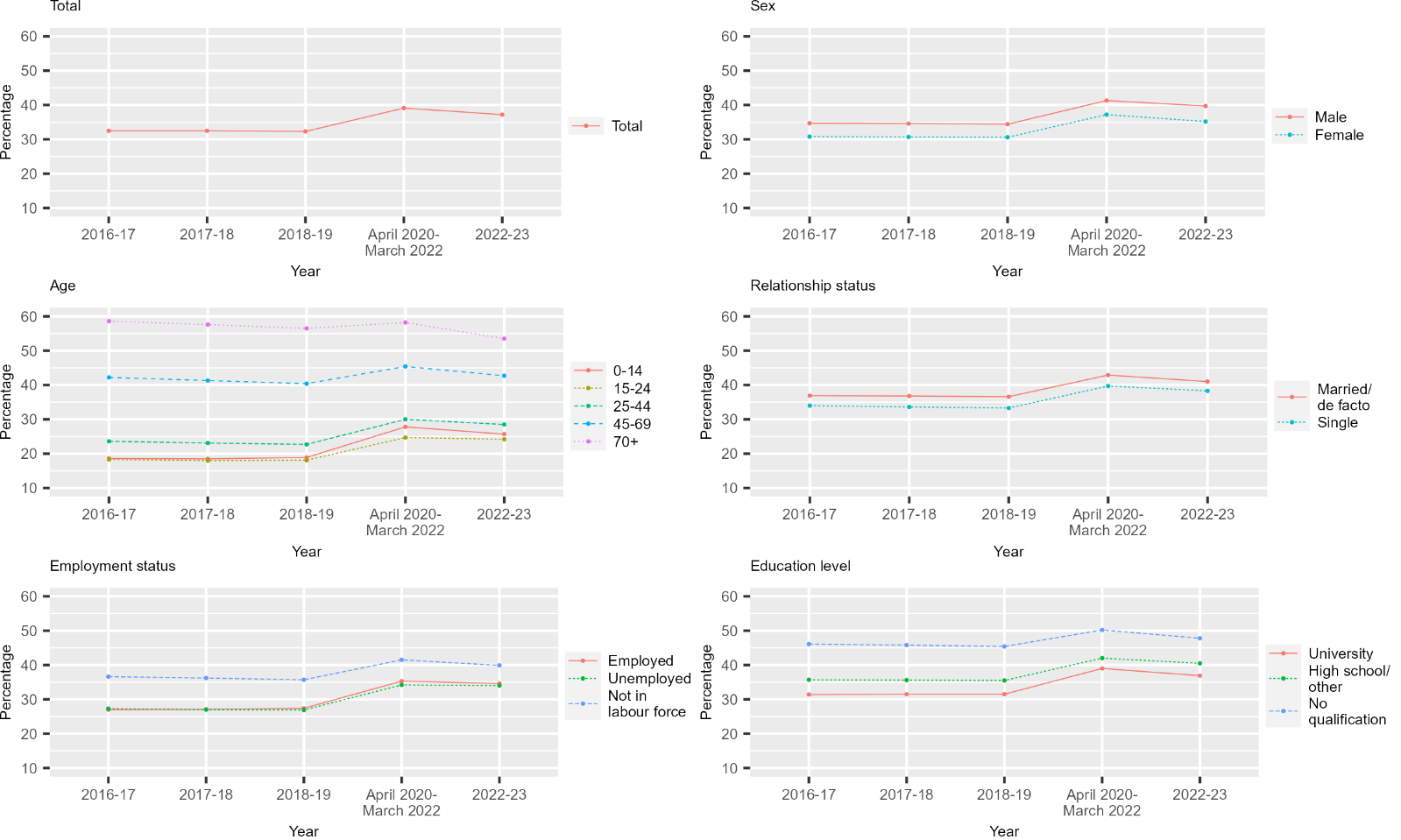

#### Supplementary Figure 4. Proportion of the Census 2016 continuity of care study population with high continuity of care (UPI score of ≥0.7) over the study period and in relation to sociodemographic characteristics.

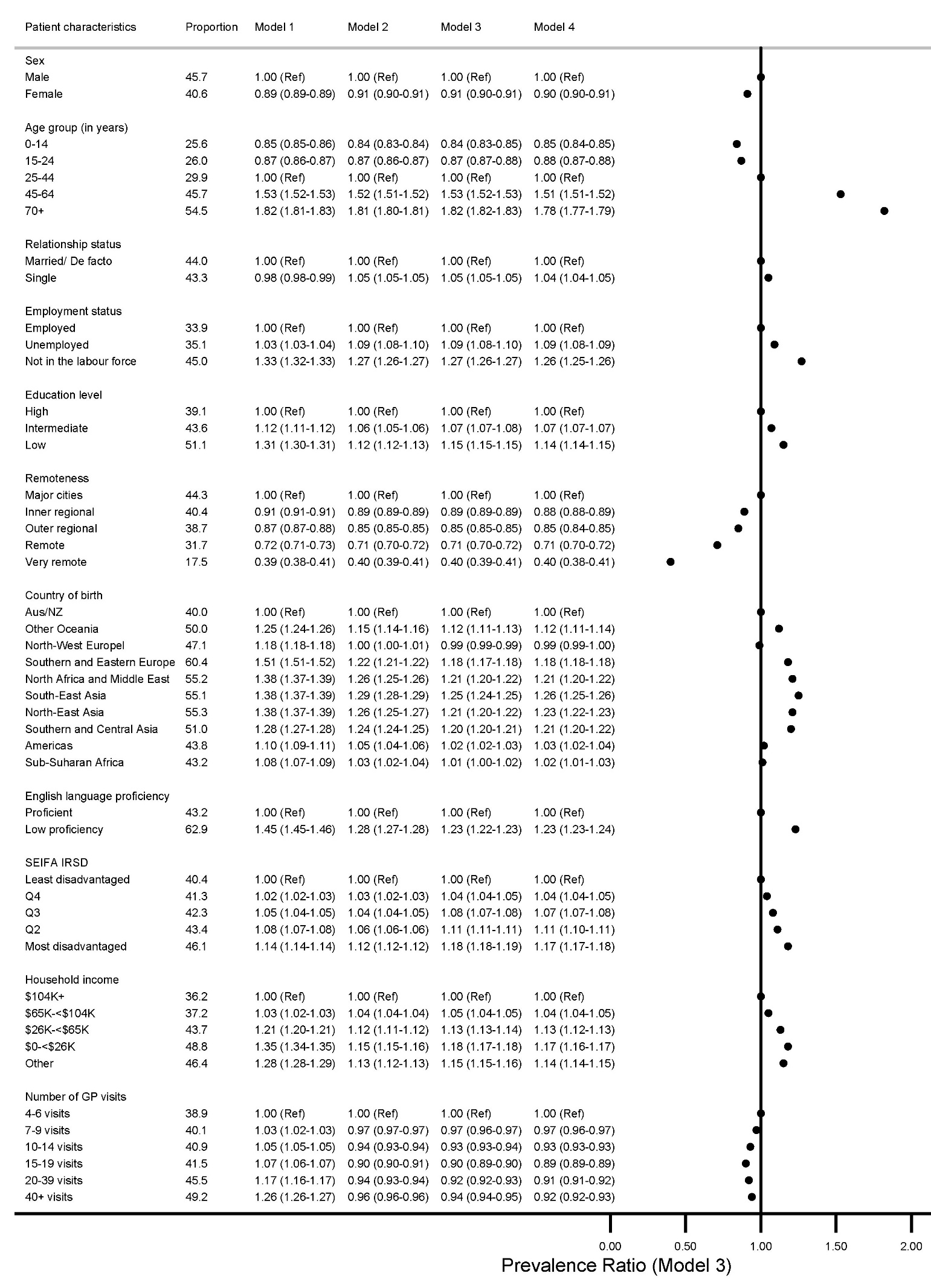
Supplementary Figure 5. Proportion of the Census 2021 continuity of care study population with at least one chronic health condition with high continuity of care and prevalence ratios (and 95% confidence intervals) describing the association between high continuity and socio-demographic, 2022-23.
